# Drug Repositioning by Merging Active Subnetworks Validated in Cancer and COVID-19

**DOI:** 10.1101/2021.05.13.21257140

**Authors:** M. Lucchetta, M. Pellegrini

## Abstract

Computational Drug Repositioning aims at ranking and selecting existing drugs for use in novel diseases or existing diseases for which these drugs were not originally designed. Using vast amounts of available omic data in digital form within an *in silico* screening has the potential for speeding up considerably the shortlisting of promising candidates in response to outbreaks of diseases such as COVID-19 for which no satisfactory cure has yet been found. We describe DrugMerge as a methodology for preclinical computational drug repositioning based on merging multiple drug rankings obtained with an ensemble of Disease Active Subnetwork construction algorithms. DrugMerge uses differential transcriptomic data from cell lines/tissues of patients affected by the disease and differential transcriptomic data from drug perturbation assays, in the context of a large gene co-expression network. Experiments with four benchmark diseases (Asthma, Rheumatoid Arthritis, Prostate Cancer, and Colorectal Cancer) demonstrate that our method detects in first position drugs in clinical use for the specified disease, in all four cases. Our method is competitive with the state-of-the-art tools such as CMAP (Connectivity Map). Application of DrugMerge to COVID-19 data found rankings with many drugs currently in clinical trials for COVID-19 in top positions, thus showing that DrugMerge is able to mimic human expert judgment.

## 1 Introduction

Among the strategies pharmacological science adopts for tackling diseases (either new or old), there are: the development of new drugs, the development of vaccines, and the repurposing of existing drugs. While the development of vaccines and novel drugs are often effective, they are also expensive and time-consuming. Thus repurposing of existing drugs is a cheaper and faster route to explore when one wishes to hedge one’s bets [Ashburn and Thor, 2004],[Nosengo, 2016].

As data about drugs and diseases accumulate in databases comprising many aspects: genomic, transcriptomic, phenotypic, clinical, and epidemiological, it becomes feasible and desirable to employ this body of accumulated knowledge within a computerized system so as to have a pre-screen *in silico* of the candidate drugs to be repurposed for a specific disease [Sam and Athri, 2019]. These shortlisted drugs are in reduced number with respect to the initial pool of drugs and thus are more amenable to the subsequent steps of the full pipeline (including *in vitro* ed *in vivo* experiments, clinical trials, and eventual deployment in clinical practice). For existing drugs often their safety profile for humans is already known, therefore once the initial *in vitro* and *in vivo* assays are positive, they can be moved directly to clinical trials of phase II or phase III (thus skipping phases 0 and I), and accelerating the drug approval lifecycle.

The recent pandemic sparked by the SARS-CoV2 virus has placed drug repurposing in the spotlight because of the pressure on the public health systems to find cures for acute cases of patients infected by SARS-CoV2. At the moment of writing, remdesivir has been approved by regulatory bodies to treat acute COVID-19 cases needing hospitalization [Rubin et al., 2020]. No other drug or treatment has passed all phases of clinical trials leading to full approval by the Federal Drug Administration (FDA) or the European Medicines Agency (EMA).

Initial drug investigations on COVID-19 were based on the accumulated experience of the efficacy of drugs *in vitro* and *in vivo* experiments for SARS-Cov, MERS, and other coronaviruses. Computerized drug repurposing however aims at a more systematic approach to candidate drug ranking and selection. Moreover, while having accrued data from other viruses belonging to the same family is a bonus, we should not always rely on having such data handy.

In a longer perspective, both the possible variations in the behavior of SARS-CoV2, due to genetic mutations, or the emergence of new human viral diseases due to cross-species transmission, imply that an easy to use, accurate, and fast pre-screening and ranking of repurposable drugs aimed at countering any new viral threats is needed. Our study is a contribution in this direction.

Network-based drug repurposing is a recent new approach to drug repurposing that may increase the effectiveness of drug repurposing pipelines. In particular network-based drug repurposing can merge many ‘omics’ data sets in a unified framework, thus such approach may be more robust and capable of including systemic biological effects [Barabasi and Oltvai, 2004], [Barabási et al., 2011], [Cheng et al., 2019], and [Gysi et al., 2020].

In [Lucchetta, 2020] we established Core&Peel as a competitive method for building disease active subnetworks compared to several state-of-the-art algorithms in several known benchmark diseases. Moreover, we applied Core&Peel and the other algorithms to build a collection of active subnetworks from gene expression data of human tissues infected by COVID-19. Subsequent pathway enrichment analysis revealed that in a COVID-19 infection are activated pathways similar to those activated in other diseases (mostly viral infections). These observations had implications for the choice of existing drugs likely to have an impact on COVID-19 to be short-listed for *in vitro* experiments and eventually go on to clinical trials. Although often the connection between a disease-related pathway and a repurposable drug was obvious in the specific context, sometimes the connection was hard to establish since, for example, for a relevant disease-related pathway too many drugs are known to be effective. Moreover, there was no quantitative way to prioritize the potentially repurposable drugs obtained from this type of qualitative analysis.

In this paper, building upon [Lucchetta, 2020], we describe a systematic way to rank repurposable drugs for a specific disease, so as to take advantage of multiple active subnetwork algorithms and multiple gene expression data sets. We use known clinically approved drugs for benchmark diseases, and trial drugs for COVID-19 as golden standard. We show that our method has better measurable performance than C-map [Lamb et al., 2006], a widely used drug ranking portal, as well as COVID-19 specific drug rankings in [Taguchi and Turki, 2020] and [Mousavi et al., 2020]. The quality of our ranking results on COVID-19 is arguably comparable to those in [Gysi et al., 2020] and [Zhou et al., 2020b], which use however quite different strategies. The whole workflow of our method is shown in Figure 1.

**Figure 1:**
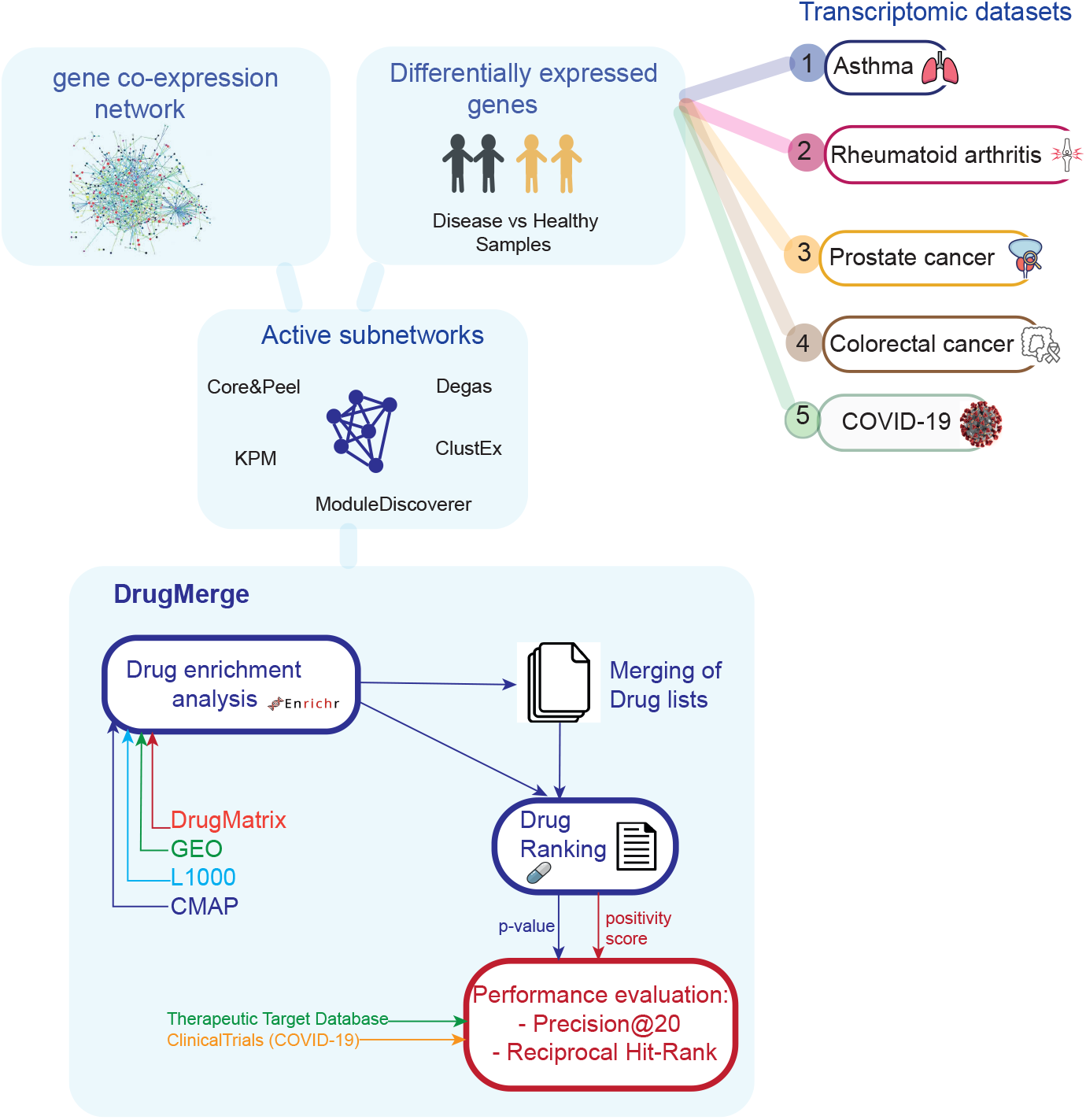
Overview of the workflow. The first part of the figure shows how we generated the data inputs of DrugMerge. These data have been created in [Lucchetta, 2020], and we reported them here for sake of completeness. We used several transcriptomic data to identify differentially expressed genes and the gene co-expression network to find one active subnetwork for each method used (Core&Peel, Modulediscoverer, Degas, ClustEx, and KeyPathwayMiner). For each of them, we performed the drug enrichment analysis on four drug perturbation databases: DrugMatrix, GEO, L1000, and CMAP. After that, we ranked the drugs according to the p-value or the positivity score. We also merged more drug lists, which come from different algorithms for the same disease and we applied the same ranking strategy. Finally, we evaluated the DrugMerge performance through the Precison@20 and the Reciprocal Hit-Rank using the golden-standard drugs, collected in the Therapeutic Target Database, and ClinicalTrials for COVID-19.

Ruiz et al. [Ruiz et al., 2020] observe that “…a drug’s effectiveness can often be attributed to targeting genes that are distinct from disease-associated genes but that affect the same functional pathways.”, in contrast to “… existing approaches assume that, for a drug to treat a disease, the proteins targeted by the drug need to be close to or even need to coincide with the disease-perturbed proteins.” Ruiz et al. thus build a multi-layered interaction graph comprising proteins, drugs, diseases, and functional pathways (extracted from the GO repository), and devise a random-walk diffusion algorithm to connect the drug nodes and the disease nodes of the model through paths that include the functional nodes of the graph.

Our approach is intermediate between the functional-based method advocated by Ruiz et al. [Ruiz et al., 2020] and protein proximity based methods (e.g. [Gysi et al., 2020]).

We do not represent GO pathways as functional nodes in the model, instead, we rely on a large and comprehensive gene co-expression network to provide the functional association in an implicit way. Dense subgraphs of the co-expression network will be formed by genes that have a coherent pattern of being up/down expressed, and thus are likely to be involved in specific cellular processes, either directly, by expressing binding proteins, or indirectly through cascading regulatory action chains, possibly involving also ncRNA and other regulatory elements not explicitly modelled. All algorithms used in this paper to build Active Network will take advantage of the dense neighborhood of the co-expression network, either directly (e.g. Core&Peel, ModuleDiscoverer) or indirectly via the augmented local graph connectivity (KPM, Degas, ClustEx). In contrast with the proximity-based methods, we do not define any ad hoc explicit distance function over a network, since the Active Network algorithms already provide a clear-cut demarcation for the part of the global gene network onto which a perturbation should act, thus we can use directly indices for measuring module enrichment or incidence of the drug’s DEG onto the disease’s Active Network.

We follow a network-based approach to drug repositioning and we focus mainly on the human transcriptional response to the disease, rather than the interaction between the disease agent (e.g. a virus) and the transcriptional response to it. In the case of COVID-19, also it is known that an over-response by the immune system is often the main cause of death in acute cases [Moore and June, 2020], [Laing et al., 2020], [Li et al., 2020] [de la Rica et al., 2020], [Tang et al., 2020], [Cron, 2020], [Hojyo et al., 2020], [Dorward et al., 2020], [Merad and Martin, 2020], and [Fajgenbaum and June, 2020].

Our strategy for COVID-19 is thus to prioritize drugs that act on a disease active subnetwork embedded into a global human gene co-expression network, to counter the virus’ global effects. This approach is complementary to a different one that aims at drugs interfering with the interactions of the host-proteins with the viral-protein. While the former approach aims at controlling the global transcriptional response of human tissues to the virus, the latter favors drugs blocking key aspects of the virus actions in entering the host cells, and activating the cells’ biochemical machinery for reproduction.

This paper is organized as follows. Section 2 reports the main results of our study on ranking for drug repositioning in four benchmark diseases and COVID-19. Section 3 describes in detail the algorithmic pipeline for the proposed DrugMerge method. Section 4 covers several strong points, current limitations, and perspective improvements of DrugMerge. Section 5 briefly gives an overview of the literature on computational drug repositioning, with more in-depth descriptions of previous methods more relevant for the development of DrugMerge. In Section 6 we make final comments on the potential of computational drug repositioning for addressing future health needs. In appendix A we give a brief synopsis of the supplementary tables and files.

## 2 Results

We compute as main quality measures the reciprocal hit ranking (RHR) and the precision at 20 (precision@20) of a ranking with respect to a golden standard (the TTD records for four benchmark diseases, and the trials.gov records for COVID-19). Each measure is then normalized via a z-score, and corresponding p-value, referred to the distribution of quality measures given by taking repeatedly random permutations of the input drug data set.

The results are shown in Figures from 2 to 9 and Supplementary Tables from S1 to S8. For DrugMerge we report only significant results at p-value ≤ 0.05 for at least one of the two quality measures. The DrugMerge method could attain significant results for all four benchmark diseases (asthma, rheumatoid arthritis, colorectal cancer, and prostate cancer) on at least one Drug Database (GEO, DrugMatrix, CMAP, L1000), and often on more than one. Moreover, as reported in the actual rankings in Supplementary Table S7, for the four benchmark data sets the reported drug ranking always has a drug in clinical use at the top of the list for the chosen Drug data set (attaining the best p-value). In Figure 6 we compare, using the p-value, the performance of DrugMerge versus the CMAP algorithm [Lamb et al., 2006] on the four benchmark diseases. DrugMerge has always a better performance in terms of p-value, even when the same CMAP drug database is used also by DrugMerge. Next we discuss in detail results for single diseases.

### 2.1 DrugMerge results on Asthma

Figure 2 and Supplementary Table S1 show that DrugMerge on the GEO Drug dataset finds one clinically relevant drug (prednisolone) in first position both when all drugs are used and when only FDA approved drugs are used. The precision@20 is not statistically significant, but the RHR of this single hit is very significant. A closer look at the GEO data and at the TTD records reveals that prednisolone is the only drug from TTD present in the GEO dataset. The initial list of differentially expressed genes is measured from a cohort of patients with acute asthma with respect to healthy patients, and interestingly the hit drug prednisolone is used specifically for acute asthma^1^. The DrugMerge algorithm does not find any hit in the L1000 drug dataset. An examination of the L1000 data shows that only 5 asthma-related drugs as reported by TTD are in the L1000 data (budesonide, fluticasone propionate, mometasone furoate, beclomethasone, and colforsin) which are used to treat chronic asthma or mild/moderate persistent asthma. Similar considerations hold for the TTD drugs present in CMAP Drug data. Thus DrugMerge appears to be able to differentiate the acute vs chronic forms of the asthma disease and rank higher drugs that are fit for the subclass of asthma patients assessed in the data set (acute asthma patients).

**Figure 2:**
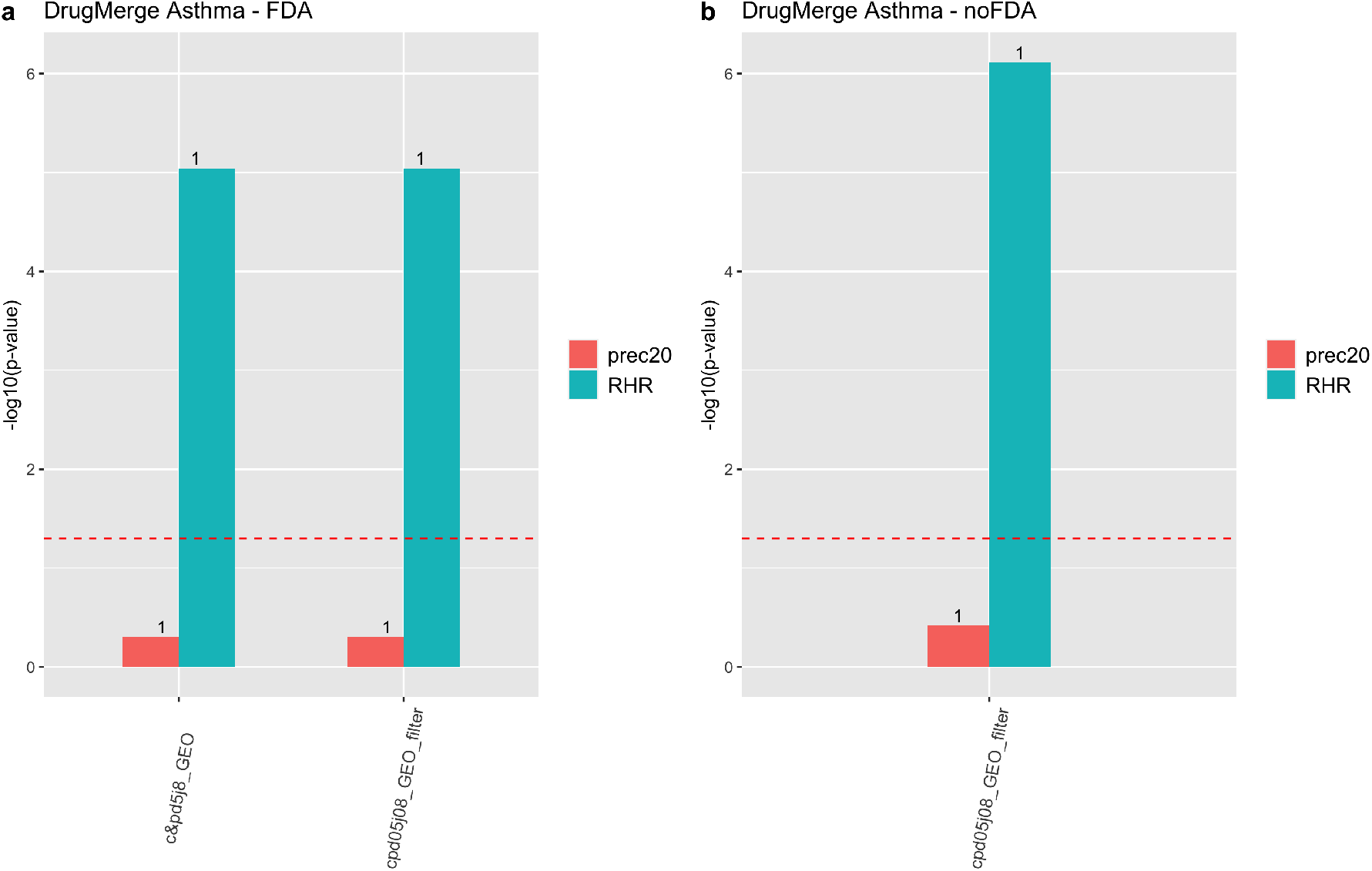
DrugMerge performance on Asthma data. The bars represent the −*log*_10_(*pvalue*) with respect to the precision@20 (red), and the RHR (light blu). On the x-axis, different algorithms or combinations of them are shown. The numbers on the top of the bars show the absolute values of precision@20 or RHR. The dotted red line represents the limit of significance (−*log*_10_(0.05)). All the bars above the dotted line show a significant p-value. The a) panel represents the DrugMerge performance when only FDA-approved drugs are considered; b) when all drugs (without any FDA filtering) are considered. The plot has been generated by the *ggplot2* [Wickham et al., 2016] R package.

Among the top positions in the ranking reported in Supplementary File S7, we find drugs with known effects in asthma patients or animal models of asthma, and one of these went into clinical trial stage: deferasirox [Sompornrattanaphan et al., 2020], rosiglitazone [Lee et al., 2016], valproic acid^2^, dexamethasone [Sarkari, 2007], celecoxib [Martin-Garcia et al., 2003], and tamoxifen [Mansilla et al., 2020].

Rosiglitazone has been tested on a murine model of chronic asthma [Lee et al., 2016], suggesting that its intranasal administration can prevent air way inflammation. A comparison study between dexamethasone versus hydrocortisone in severe acute pediatric asthma [Sarkari, 2007] showed that the mean length of hospitalization in children receiving dexamethasone was significantly shorter than those receiving hydrocortisone. Celecoxib is a COX-2 inhibitor and nonsteroidal anti-inflammatory drug.

A study of 33 asthma patients [Martin-Garcia et al., 2003] demonstrated that celecoxib is a suitable drug in aspirin-induced and/or nonsteroidal anti-inflammatory drug-induced asthma patients. Tamoxifen is an estrogen receptor modulator mainly used for the treatment of breast cancer, but it has also been reported to have anti-inflammatory activity. A recent study [Mansilla et al., 2020] has tested tamoxifen on three different mouse models, showing that tamoxifen can reduce inflammatory infiltration of neutrophils in the airways.

For the remaining drugs in the top positions, we focus on their mechanism of action and we search in literature if this can have relevance in asthma. Using this criterion, we find captopril, letrozole, decitabine, imatinib, and probucol as relevant drugs. Captopril belongs to the class of drugs known as angiotensin-converting enzyme (ACE) inhibitors and is used primarily to lower high blood pressure (hypertension). It has been proved that asthmatic subjects with comorbid hypertension display evidence of enhanced asthma morbidity [Christiansen et al., 2016].

Letrozole inhibits aromatase, an enzyme that catalyzes the synthesis of estrogen. Asthma prevalence and severity are greater in women than in men, suggesting this is in part related to female steroid sex hormones, such as estrogen. Estrogen receptors are found on numerous immunoregulatory cells and estrogen’s actions skew immune responses toward allergy [Bonds and Midoro-Horiuti, 2013].

Decitabine is a hypomethylating agent and is used to treat myelodysplastic syndromes and acute myeloid leukemia. Yang et al. [Yang et al., 2015] demonstrated that DNA methylation in specific gene loci are associated with asthma and suggest that epigenetic changes might play a role in establishing the immune phenotype associated with childhood asthma.

Imatinib is a tyrosine kinase inhibitor. Anti-inflammatory effects of tyrosine kinase inhibitors have been reported in animal models of allergic asthma, suggesting that this kind of drug can be a very attractive strategy for the treatment of asthma [Wong and Leong, 2004].

Finally, probucol lowers the level of cholesterol through the inhibition of cholesterol synthesis and deletion of cholesterol absorption. Ramaraju et al. [Ramaraju et al., 2013] found a modest but significant association between higher levels of serum cholesterol and asthma.

### 2.2 DrugMerge results on Rheumatoid Arthritis (RA)

Figures 3 and Supplementary Table S2 show that DrugMerge finds significant rankings in several drug databases (L1000, DrugMatrix, GEO), with GEO attaining the best p-value, using several algorithms, both when all drugs are considered or when only FDA-approved drugs are considered. For the GEO dataset Supplementary Table S7, we find in first position the clinically relevant drug dexamethasone. Other drugs in use according to the TTD records that appear in the top twenty positions are: imatinib, methylprednisolone, and prednisolone.

**Figure 3:**
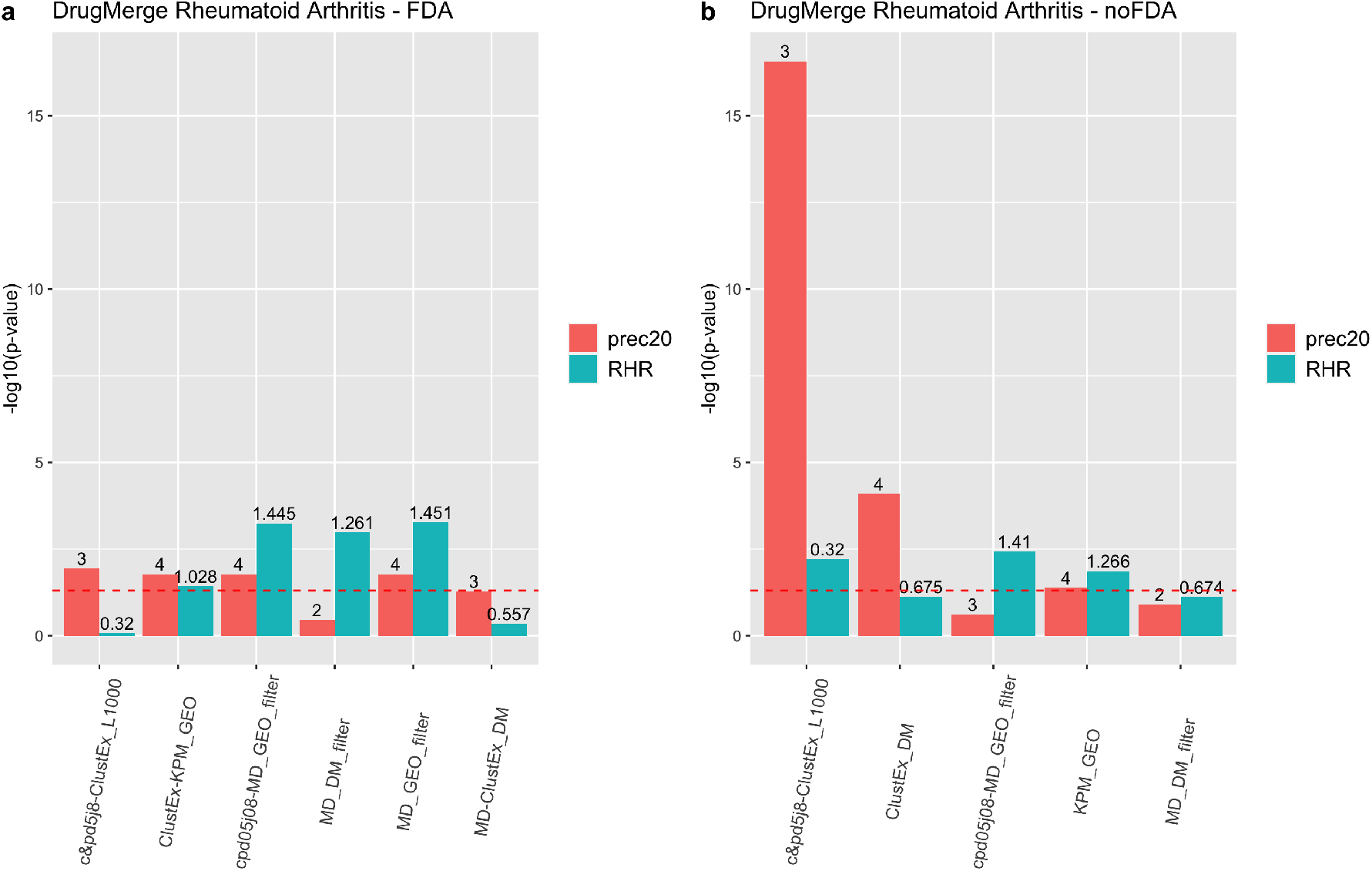
DrugMerge performance on Rheumatoid Arthritis data. The bars represent the −*log*_10_(*pvalue*) with respect to the precision@20 (red), and the RHR (light blu). On the x-axis, different algorithms or combinations of them are shown. The numbers on the top of the bars show the absolute values of precision@20 or RHR. The dotted red line represents the limit of significance (−*log*_10_(0.05)). All the bars above the dotted line show a significant p-value. The a) panel represents the DrugMerge performance when only FDA-approved drugs are considered; b) when all drugs (without any FDA filtering) are considered. The plot has been generated by the *ggplot2* [Wickham et al., 2016] R package.

Among the top positions in the ranking, we find drugs with known effects in rheumatoid arthritis patients or animal models of rheumatoid arthritis, and one of these went into clinical trial stage: rosiglitazone^3^, pioglitazone [Suke et al., 2013], estradiol [Romo-García et al., 2020], captopril [Martin et al., 1984], vitamin c [Carr et al., 2015], baclofen [Huang et al., 2015], deferasirox [Polson et al., 1986], decitabine [Petralia et al., 2019], sirolimus [Wen et al., 2019], hydrocortisone[Boland, 1952], paclitaxel [Kurose et al., 2001], and ethinylestradiol [Subramanian et al., 2005].

Suke et al. [Suke et al., 2013] studied the effects of combined pioglitazone and prednisolone on adjuvant-induced arthritis in rats. This study suggested that the combination of these two drugs was effective in modulating the inflammatory response and suppress arthritis progression.

Estradiol is an estrogen steroid female hormone, and estrogens have a direct action upon the immune system. The role of the estrogens in rheumatoid arthritis has been studied here [Romo-García et al., 2020].

Captopril is prescribed for hypertension but it has immunosuppressant activity, as well. Therefore, captopril was considered a potential slow-acting drug for treating rheumatoid arthritis as demonstrated in [Martin et al., 1984].

Vitamin C is a vitamin found in various foods and it is important for immune system function. It has also been studied the role of the vitamin c in treating pain [Carr and McCall, 2017], in particular, an administration of high-dose vitamin C in patients with rheumatoid arthritis showed a complete decrease in pain [Carr et al., 2015].

Baclofen is a medication used to treat muscle spasticity. Huang et al. [Huang et al., 2015] investigated the effects of baclofen in murine collagen-induced arthritis, proving that baclofen alleviated the clinical development of arthritis.

Decitabine has already been found in asthma analysis and as mentioned before, it inhibits DNA methylation. Petralia et al. [Petralia et al., 2019] have studied the effect of decitabine in a murine model of rheumatoid arthritis and have demonstrated that decitabine administration was associated with a significant improvement of the clinical condition.

Sirolimus, also known as rapamycin, has immunosuppressant functions and is used to prevent rejection in organ transplants. Wen et al. [Wen et al., 2019] studied the safety, tolerance, and efficacy of sirolimus in patients with active RA treated with low-dose sirolimus combined with original therapy. They showed that this therapy alleviates clinical symptoms and decreases the immunosuppressive applications in patients with active RA.

Hydrocortisone is a treatment for acute episodes of rheumatic disorders, including rheumatoid arthritis ^4^.

Paclitaxel is an anticancer agent and is classified as a microtubule-stabilizing agent. Kurose et al. [Kurose et al., 2001] studied the effects of paclitaxel on cultured synovial cells from patients with rheumatoid arthritis. The data suggest paclitaxel as a possible therapy for RA.

Ethinylestradiol is an active estrogen and component of birth control pills. Subramaniam et al. [Subramanian et al., 2005] studied the effectiveness of ethinylestradiol in treating collagen-induced arthritis mice, noticing a decreased proliferation and secretion of pro-inflammatory factors.

Focusing on the action mechanism of the remaining drugs, we find some links with rheumatoid arthritis, such as amoxicillin [Ogrendik, 2014], and niacin [Peclat et al., 2020].

Amoxicillin is an antibiotic used to treat several bacterial infections. Since the 1930s, RA has been treated with antibiotics and there have been several reports in the literature indicating that periodontal pathogens are a possible cause of the disease [Ogrendik, 2014].

Niacin, more commonly known as vitamin B3, is a precursor of the coenzymes nicotinamide-adenine dinucleotide (NAD+). Recent studies have identified potential therapeutic approaches for boosting NAD+ to treat rheumatologic diseases, including rheumatoid arthritis. In particular, they focused on the enzymatic activity of CD38, one of the main enzymes in NAD+ catabolism [Peclat et al., 2020].

### 2.3 DrugMerge results on Colorectal Cancer

Figures 4 and Supplementary Table S3 show that DrugMerge finds significant rankings in several drug databases (L1000, CMAP, GEO), using several algorithms, both when all drugs are considered or when only FDA-approved drugs are considered. For the L1000 dataset Supplementary Table S7, we find in the first position the clinically relevant drug erlotinib. Other drugs in clinical use according to the TTD records that appear in the the top twenty positions are: lapatinib and sunitinib. Among the top positions in the ranking, we find drugs with known effects in colorectal cancer patients, related animal models, or cell lines, and one of these went into clinical trial stage: mitoxantrone [Cornelia et al., 1991], sorafenib [Kacan et al., 2016], sutent (sunitinib) ^5^, dasatinib [Scott et al., 2017], sirolimus (rapamycin) [Wagner et al., 2009], azacitidine [Azad et al., 2017], imatinib [Samei et al., 2016], tamoxifen [Kuruppu et al., 1998], doxorubicin [Lee et al., 2017], naltrexone [Ma et al., 2020], and digoxin [Hou et al., 2020].

**Figure 4:**
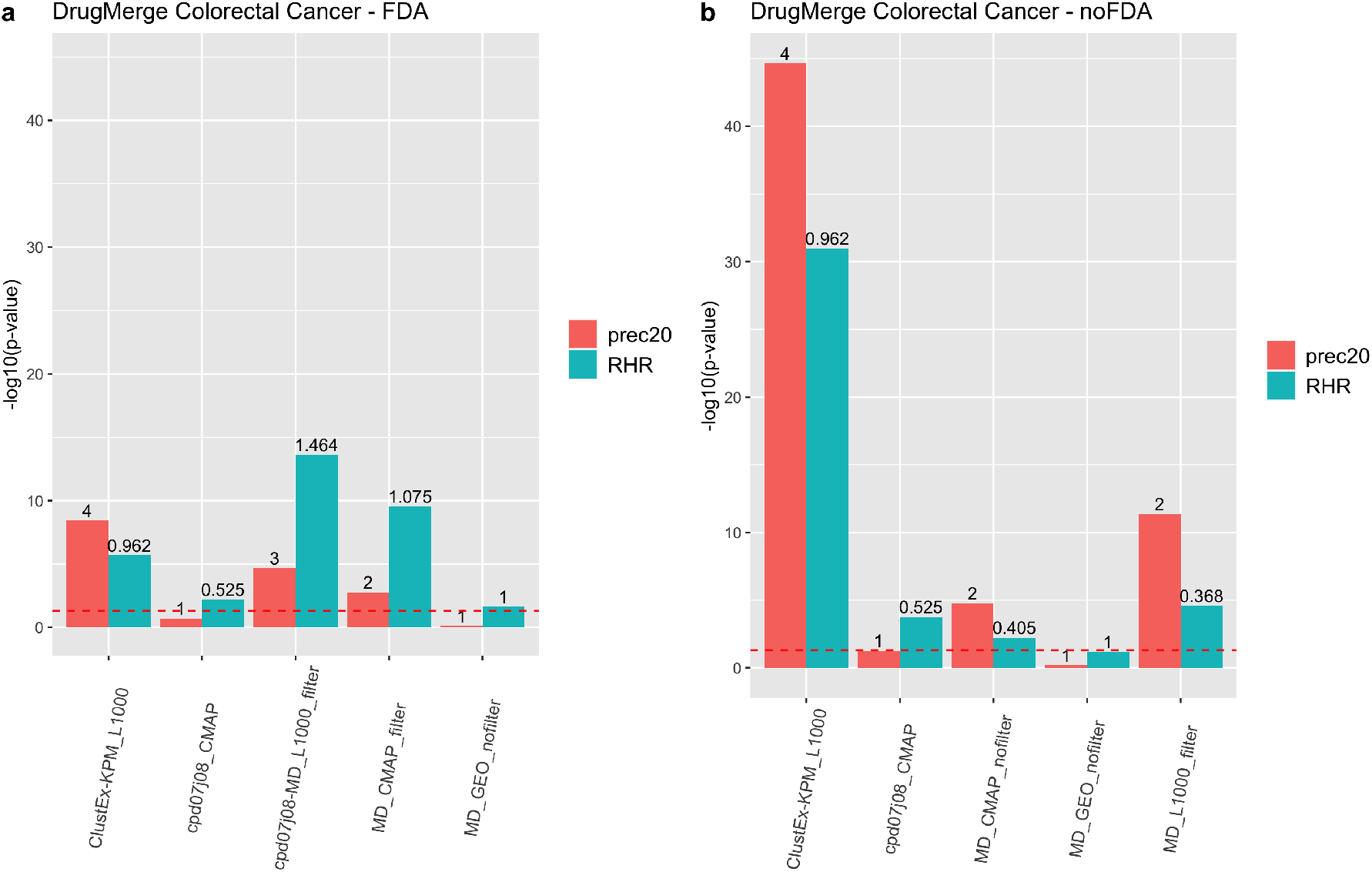
DrugMerge performance on Colorectal Cancer data. The bars represent the −*log*_10_(*pvalue*) with respect to the precision@20 (red), and the RHR (light blu). On the x-axis, different algorithms or combinations of them are shown. The numbers on the top of the bars show the absolute values of precision@20 or RHR. The dotted red line represents the limit of significance (−*log*_10_(0.05)). All the bars above the line show a significant p-value. The a) panel represents the DrugMerge performance when only FDA-approved drugs are considered; b) when all drugs (without any FDA filtering) are considered. The plot has been generated by the *ggplot2* [Wickham et al., 2016] R package.

Mitoxantrone has been tested in 13 patients with advanced or metastatic colon carcinoma and the authors observed that mitoxantrone was moderately effective [Cornelia et al., 1991].

Sorafenib is a kinase inhibitor drug approved for the treatment of some cancers. In [Kacan et al., 2016], the authors showed that sorafenib has a strong antitumor and antiangiogenic effect on the colorectal cancer cell line.

Scott et al. [Scott et al., 2017] demonstrated that dasatinib had significant anti-proliferative activity in a subset of a colorectal cancer cell line, even if dasatinib is currently being studied in combination with chemotherapy, as its use as a single agent appears limited.

Wagner et al. [Wagner et al., 2009] studied the effects of rapamycin in mice and showed that it was able to suppress advanced stage colorectal cancer.

Azacitidine has been tested in combination with entinostat in metastatic colorectal cancer patients, resulting in a tolerable therapy [Azad et al., 2017].

Imatinib is an oral chemotherapy medication used to treat cancer and it has been proved that it inhibits proliferation of colon cancer cells [Samei et al., 2016].

Kuruppo et al [Kuruppu et al., 1998] demonstrated that tamoxifen has a potent inhibitory action on colorectal cancer metastases in a murine model.

Doxorubicin is a chemotherapy medication used to treat cancer. In [Lee et al., 2017] the authors propose doxorubicin-loaded oligonucleotides attached to gold nanoparticles as a drug delivery system for cancer chemotherapy. They tested this approach in colon cancer cell lines, demonstrating the drug’s efficacies such as *in vitro* cytotoxicity.

Naltrexone is primarily used to manage alcohol or opioid dependence. Ma et al. [Ma et al., 2020] explored the inhibitory effect of low-dose naltrexone on colorectal cancer progression *in vivo* and *in vitro*.

Digoxin is used to treat various heart conditions such as atrial fibrillation, and heart failure. A recent study [Hou et al., 2020] has examined the effects of digoxin on two kinds of colorectal cancer cells. Their findings suggest that digoxin has the potential to be applied as an antitumor drug via inhibiting proliferation and metastasis.

For the remaining drugs in the top twenty positions, we focus on their action mechanism, and we find some links with colorectal cancer: epirubicin [Coss et al., 2009], everolimus [Thiem et al., 2013], itavastatin [Li et al., 2021], and dactinomycin [Xu et al., 2020].

Epirubicin is a chemotherapy agent, intercalating into DNA and inhibiting topoisomerase II, thereby inhibiting DNA replication. In [Coss et al., 2009] the authors demonstrate topoisomerase IIa gene and protein alterations in colorectal cancer and show that increased expression is associated with pathologically advanced disease.

Everolimus is a medication used as an immunosuppressant and in the treatment of several tumors. Its main mechanism of action is the mTOR inhibition. Thiem et al. [Thiem et al., 2013] demonstrated that the mTORC1 inhibition reduces the inflammation associated with the gastrointestinal tumor. Itavastatin (or pitavastatin) belongs to the class of statins, which are inhibitors of HMG-CoA reductase, the enzyme that catalyzes the first step of cholesterol synthesis. A meta-analysis work [Li et al., 2021] showed that statin use is a protective factor for colorectal cancer prognosis, and it might be significantly associated with lower overall mortality.

Finally, dactinomycin is a chemotherapy medication and can inhibit transcription. Recent studies have revealed crucial roles of transcription regulation in colorectal cancer development. In particular, deregulation of transcription factors is pretty frequent, and drastic changes in gene expression profiles play fundamental roles in the multistep process of tumorigenesis [Xu et al., 2020].

### 2.4 DrugMerge results on Prostate Cancer

Figures 5 and Supplementary Table S4 show that DrugMerge finds significant rankings in several drug databases (L1000, GEO) for prostate cancer, using several algorithms, both when all drugs are considered or when only FDA approved drugs are considered. For the L1000 data set Supplementary Table S7, we find in the first position the clinically relevant drug mitoxantrone. Other drugs in clinical use according to the TTD records that appear in the top twenty positions are: dasatinib, decitabine, menadione, and doxorubicin hydrochloride.

**Figure 5:**
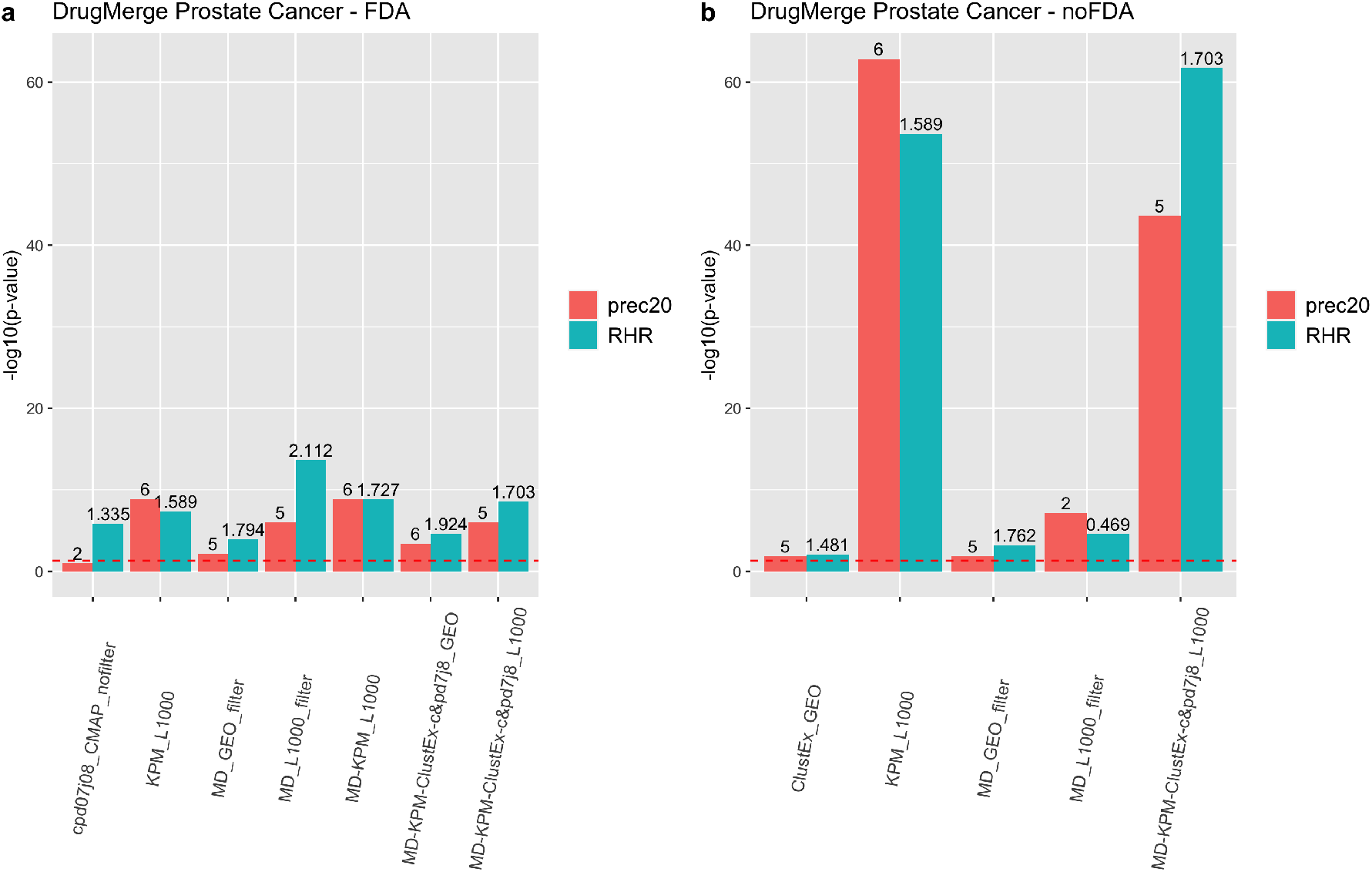
DrugMerge performance on Prostate Cancer data. The bars represent the −*log*_10_(*pvalue*) with respect to the precision@20 (red), and the RHR (light blu). On the x-axis, different algorithms or combinations of them are shown. The numbers on the top of the bars show the absolute values of precision@20 or RHR. The dotted red line represents the limit of significance (−*log*_10_(0.05)). All the bars above the dotted line show a significant p-value. The a) panel represents the DrugMerge performance when only FDA-approved drugs are considered; b) when all drugs (without any FDA filtering) are considered. The plot has been generated by the *ggplot2* [Wickham et al., 2016] R package.

**Figure 6:**
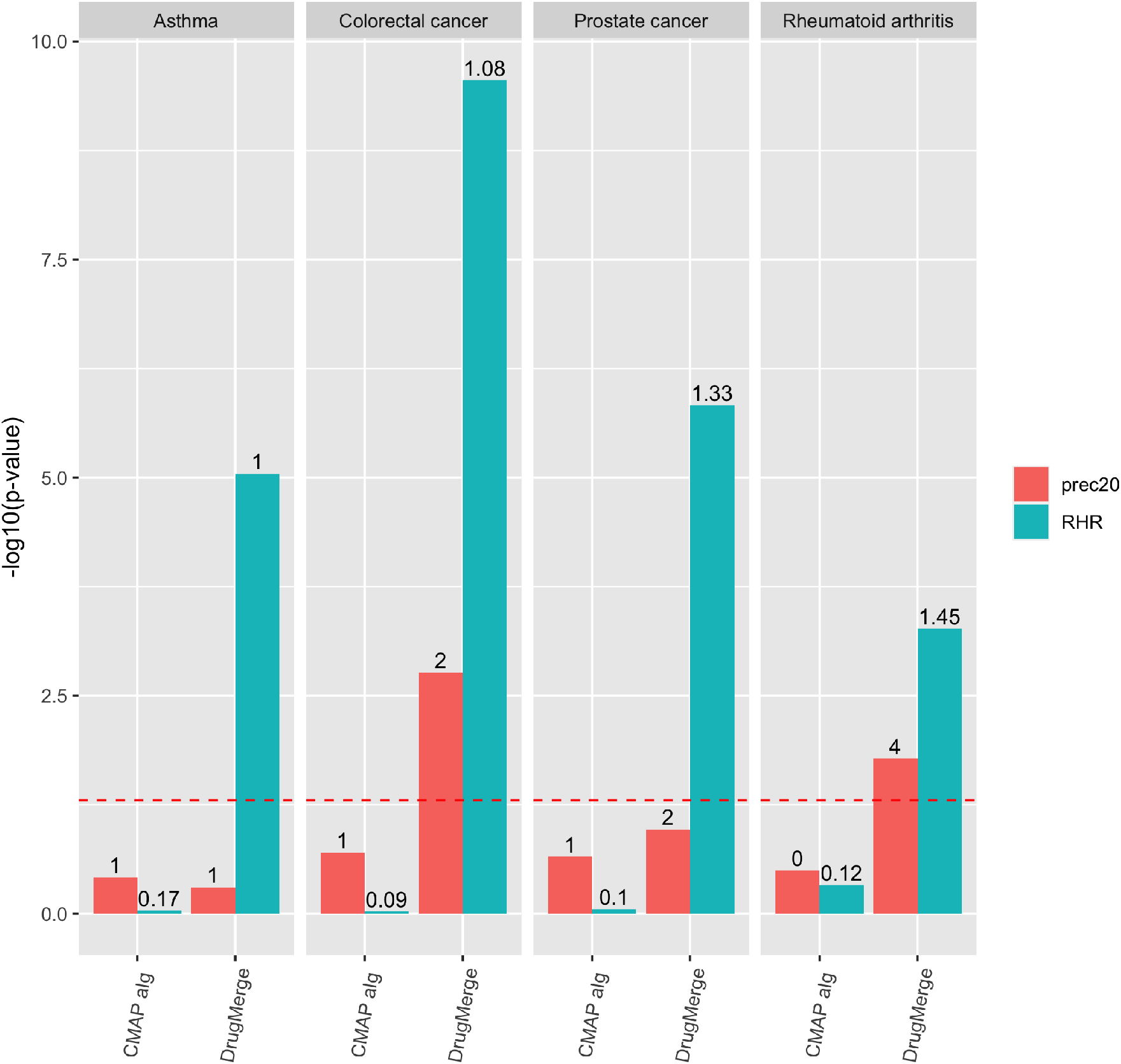
Comparison of performance of DrugMerge and the CMAP algorithm across the four benchmark diseases. For Colorectal Cancer and Prostate Cancer both methods use CMAP data. For Asthma and Rheumatoid Artrithis DrugMerge uses GEO data. The bars represent the −*log*_10_(*pvalue*) with respect to the precision@20 (red), and the RHR (light blu). The numbers on the top of the bars show the absolute values of precision@20 or RHR. The dotted red line represents the limit of significance (−*log*_10_(0.05)). All the bars above the dotted line show a significant p-value. The plot has been generated by the *ggplot2* [Wickham et al., 2016] R package.

Among the top twenty positions in the ranking we find drugs with known effects in prostate cancer patients, related animal models, or cell lines, and one of these went into clinical trial stage: erlotinib [Gravis et al., 2008], sorafenib [Zaborowska et al., 2012], lapatinib [Whang et al., 2013], sirolimus [Nandi et al., 2021], sutent (sunitinib) [Michaelson et al., 2009], azacitidine [Singal et al., 2015], everolimus [Morgan et al., 2008], gemcitabine^6^, ciclosporine [Kawahara et al., 2015], and auranofin [Liu et al., 2019].

A study of 30 patients with advanced or metastatic prostate cancer has been conducted to test the effect of erlotinib [Gravis et al., 2008], and it demonstrated an improvement in clinical benefit.

Sorafenib has been tested in patients with hormone-refractory prostate cancer [Zaborowska et al., 2012], indicating that the treatment may be associated with good outcomes in terms of overall survival.

Lapatinib is a dual tyrosine kinase inhibitor of the epidermal growth factor 1 (EGFR) and 2 (HER2). In a study of twenty-nine castration-resistant prostate cancer patients [Whang et al., 2013], lapatinib showed single-agent activity in a small subset of unselected patients.

Nandi et al. [Nandi et al., 2021] investigated the effect of sirolimus encapsulated liposomes in two different prostate cancer cell lines, and noticed an antiproliferative effect.

Sunitinib, an inhibitor of tyrosine kinase receptors, has been studied in men with castration-resistant prostate cancer [Michaelson et al., 2009], indicating that it is well tolerated and may have modest benefit in this patient population.

Azacitidine, a demethylating agent, has been tested in combination with docetaxel, and prednisone in patients with metastatic castration-resistant prostate cancer and showed to be active in this subset of population [Singal et al., 2015].

Everolimus, an mTOR inhibitor, has been assessed in mice, resulting in an inhibition of prostate cancer growth [Morgan et al., 2008].

For the remaining drugs, we focus on their action mechanism, and we find some links with prostate cancer for imatinib [Vicentini et al., 2003], epirubicin [Patra et al., 2011], and mitomycin C [Gurumurthy et al., 2001].

Imatinib is an oral chemotherapy medication and functions as a tyrosine kinase inhibitor. Vicentini et al. [Vicentini et al., 2003] investigated the effects of the epidermal growth factor receptor tyrosine kinase inhibitor on human prostatic cancer cell lines, suggesting that it may have the potential in blocking tumor growth and progression.

Epirubicin has been already found in colorectal cancer and as mentioned before, it is a topoisomerase II inhibitor. An *in vitro* cell-based assay [Patra et al., 2011] demonstrated that MHY336, a topoisomerase II inhibitor, significantly inhibited the proliferation of three prostate cancer cell lines. Finally, mitomycin C is used as a chemotherapeutic agent and it is a potent DNA crosslinker. DNA crosslinking has useful merit in chemotherapy and targeting cancerous cells for apoptosis [Deans and West, 2011]. It is known that evading cell death is one hallmark of cancer, in particular, prostate cancer cells develop multiple apoptosis blocking strategies during the various stages of tumor progression [Gurumurthy et al., 2001].

### 2.5 DrugMerge results on COVID-19

Figures 7 and 8 and Supplementary Table S5 show that DrugMerge finds significant rankings in all drug databases (L1000, CMAP, GEO, DrugMatrix), using a variety of algorithms and their combinations, both when all drugs are considered and when only FDA approved drugs are considered. For COVID-19 we have a choice of three disease data sets (cells, BALF, PBMC), four Drug datasets (L1000, CMAP, GEO, and DrugMatrix), and several algorithms and weighting functions. We proceed by first running a single algorithm on a single drug data set, collecting the configurations that give the best significant performance measures. Next, we seek improvements by merging the rankings for several algorithms with significant performance on a single disease data set. And finally, we combine the improved significant results by merging the rankings obtained by several significant algorithms on several COVID-19 disease data sets. Figures 7 and 8 and Supplementary Table S5 report the best results in each phase, while Table S5.1 report the full algorithmic settings.

**Figure 7:**
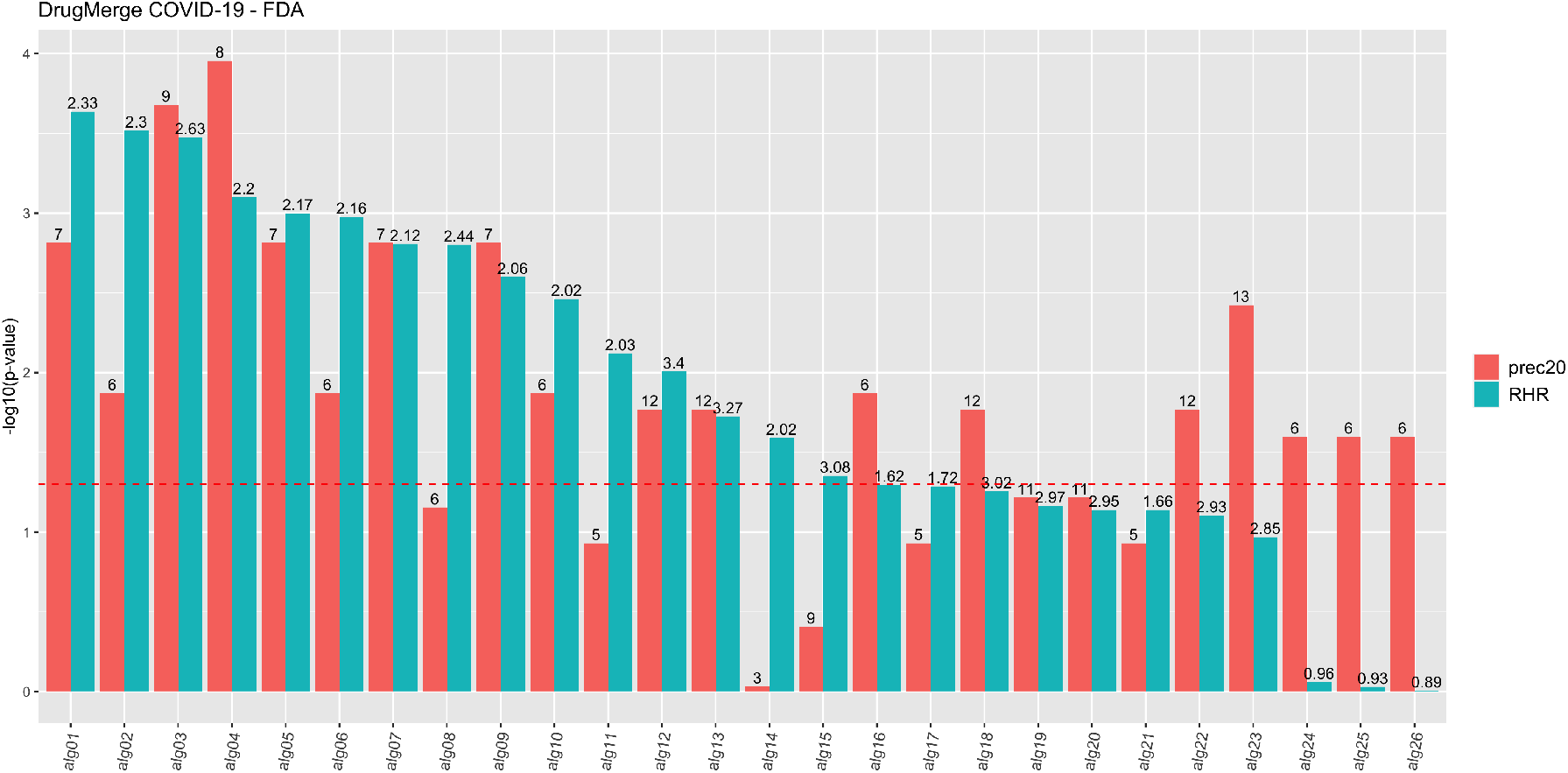
DrugMerge performance on COVID-19 data restricted to drugs with FDA approval. The bars represent the −*log*_10_(*pvalue*) with respect to the precision 20 (red), and the RHR (light blu). On the x-axis, different algorithms or combinations of them are indexed with aliases, since the complete algorithm names are too long. The mapping between them is on Table S5.1. The numbers on the top of the bars show the absolute values of precision@20 or RHR. The dotted red line represents the limit of significance (−*log*_10_(0.05)). All the bars above the dotted line show a significant p-value.

**Figure 8:**
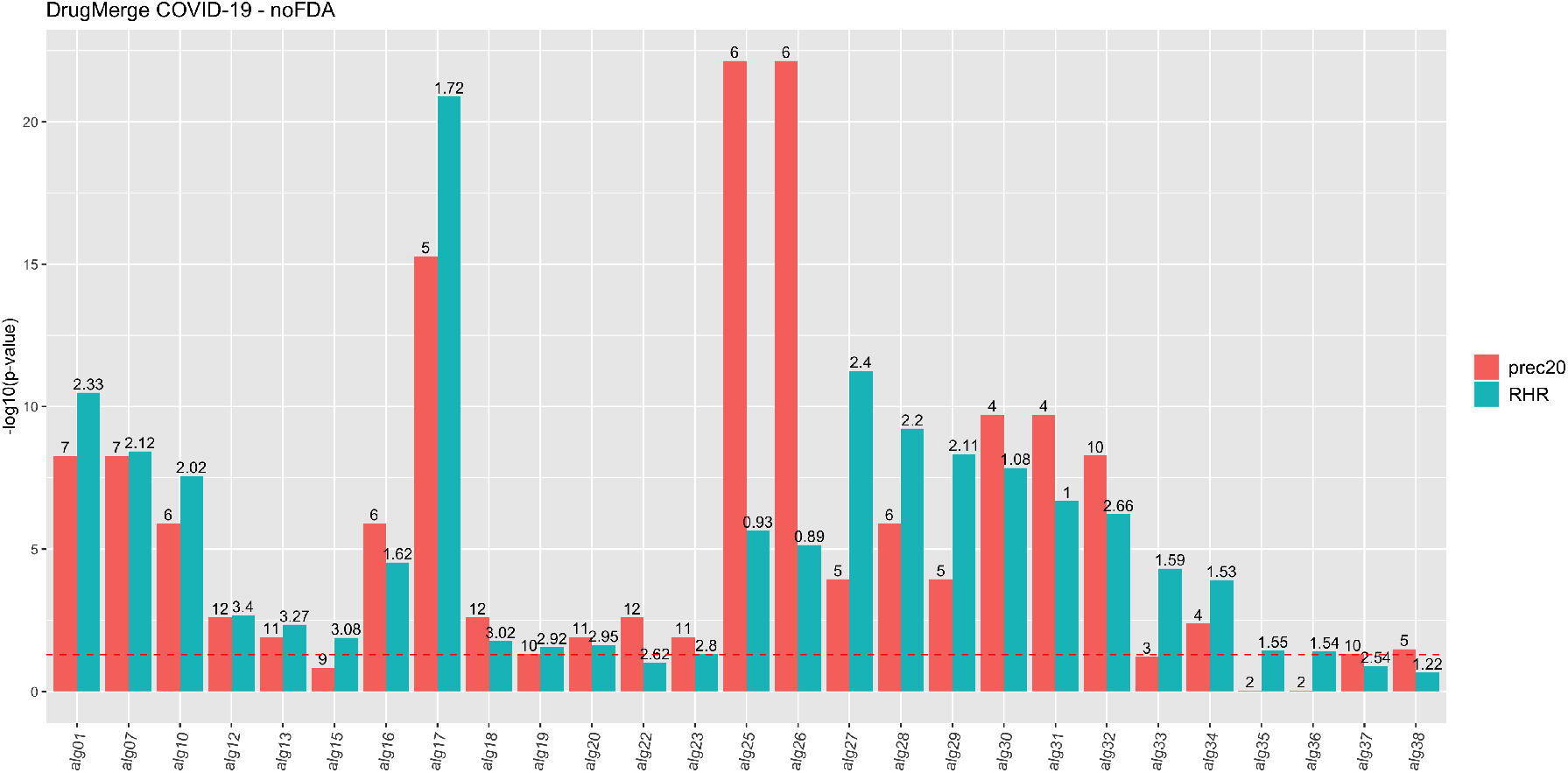
DrugMerge performance on COVID-19 data without FDA approval restriction. The bars represent the −*log*_10_(*pvalue*) with respect to the precision@20 (red), and the RHR (light blu). On the x-axis, different algorithms or combinations of them are indexed with aliases, since the complete algorithm names are too long. The mapping between them is on Table S5.1. The numbers on the top of the bars show the absolute values of precision@20 or RHR. The dotted red line represents the limit of significance (−*log*_10_(0.05)). All the bars above the dotted line show a significant p-value.

We focus now on two such rankings in Supplementary file S7 attaining the best p-values on RHR for all drugs or FDA-approved drugs only.

The highest p-value on RHR when only FDA-approved drugs are considered is attained on the CMAP Drug dataset using all three disease data sets for COVID-19 and three algorithms (ClustEx, Core&Peel, Degas), (Alg 01 in Figure 7, see Supplementary Table S5.1). The top two positions of the ranking report two drugs listed in clinical trial on www.trials.gov for COVID-19: etoposide and mefloquine. Within the first twenty positions, further 5 drugs are in clinical trials for COVID-19: colchicine, chlorpromazine, propofol, tretinoin, and ivermectin. Among the top ten positions in the ranking we find drugs with known effects animal models or cell lines or proposed for further scrutiny *in silico* screenings: thioridazine [Otręba et al., 2020], prochlorperazine [Kow and Hasan, 2020], primaquine [Sachdeva et al., 2020], fluphenazine [Nazeam et al., 2020].

The highest p-value on RHR when all drugs are considered is attained by KPM on the L1000 drug data set with the BALF COVID-19 disease data set (Alg 17 in Figure 8, see Suppl. Table S5.1). The top position of the ranking reports a drug listed in clinical trial on www.trials.gov for COVID-19: sirolimus. Within the first twenty positions further 4 drugs are in clinical trials for covid19: decitabine, tamoxifen, simvastatin, and imatinib. Among the top ten positions in the ranking we find drugs with known effects in animal models or cell lines or proposed for further scrutiny in *in silico* screenings: lapatinib [Raymonda et al., 2020], dasatinib [Weisberg et al., 2020], mitoxantrone [Lokhande et al., 2020], everolimus [Terrazzano et al., 2020], sunitinib [Wang et al., 2020b].

For the remaining drugs in the top ten positions in both lists, we determine the main mechanism of action (i.e we determine the main pathways affected by the drug having the desired pharmacological effect) and we then look for hints in the literature of relevance of these pathways for COVID-19.

In the first list, we find methylergometrine, which is a partial agonist and antagonist of serotonin, dopamine, and *α*-adrenergic receptors. Costa et al. [Costa et al., 2020] conjectured that serotonin inhibitors could have a neuroprotective effect in patients affected by COVID-19.

In the second list, we find erlotinib, stuent/sunitinib, azacitidine, and sorafenib. Erlotinib is an epidermal growth factor receptor (EGFR) inhibitor in use for the treatment of non-small cell lung cancer (NSCLC) and pancreatic cancer. Epidermal growth factor receptor (EGFR) signaling is known to be involved in the progress of viral infections [Hondermarck et al., 2020], including those caused by SARS [Venkataraman and Frieman, 2017] and SARS-Cov2 [Klann et al., 2020], thus it is a plausible target pathway to consider.

Sunitinib (Sutent) multi-targeted receptor tyrosine kinase (RTK) inhibitor used in the treatment of renal cell carcinoma (RCC) and imatinib-resistant gastrointestinal stromal tumor (GIST). Weidberg et al. [Weisberg et al., 2020] recently advocated the repurposing of kinase inhibitors for the treatment of COVID-19.

Azacitidine has been shown effective against human immunodeficiency virus (HIV) *in vitro* [Dapp et al., 2009] and human T-lymphotropic virus (HTLV) [Diamantopoulos et al., 2012]. Its action is mainly at a epigenetic level, inhibiting of DNA methyltransferase, causing hypomethylation of DNA. The role of epigenetics in COVID-19 is a growing area of research with large potential for advances in drug target identification [Pruimboom, 2020], [Schäfer and Baric, 2017], and [Chlamydas et al., 2021].

Sorafenib is a kinase inhibitor drug approved for the treatment of advanced primary renal cell carcinoma, advanced primary liver cancer, and forms of resistant advanced thyroid carcinoma. Sorafenib is a protein kinase inhibitor with activity against the Raf/Mek/Erk pathway [Liu et al., 2006]. Several authors have explored the role of MAP Kinase signalling pathway in the coronavirus family at large [Cai et al., 2007] and for SARS-cov2 in particular [Hemmat et al., 2021],[Ghasemnejad-Berenji and Pashapour, 2021], and [Zhou et al., 2020a].

In Supplementary File S8 and figure 9 we report performance measures for the proposed COVID-19 drug repurposing rankings from Taguchi et al 2020 [Taguchi and Turki, 2020], Mousavi et al 2020 [Mousavi et al., 2020], Gysi et al 2020 [Gysi et al., 2020], and Zhou et al 2020 [Zhou et al., 2020b]. For Taguchi et al 2020, and Mousavi et al 2020 the drug data sets used are overlapping with ours thus it is possible to compute the significance of the performance measures on a common basis.

**Figure 9:**
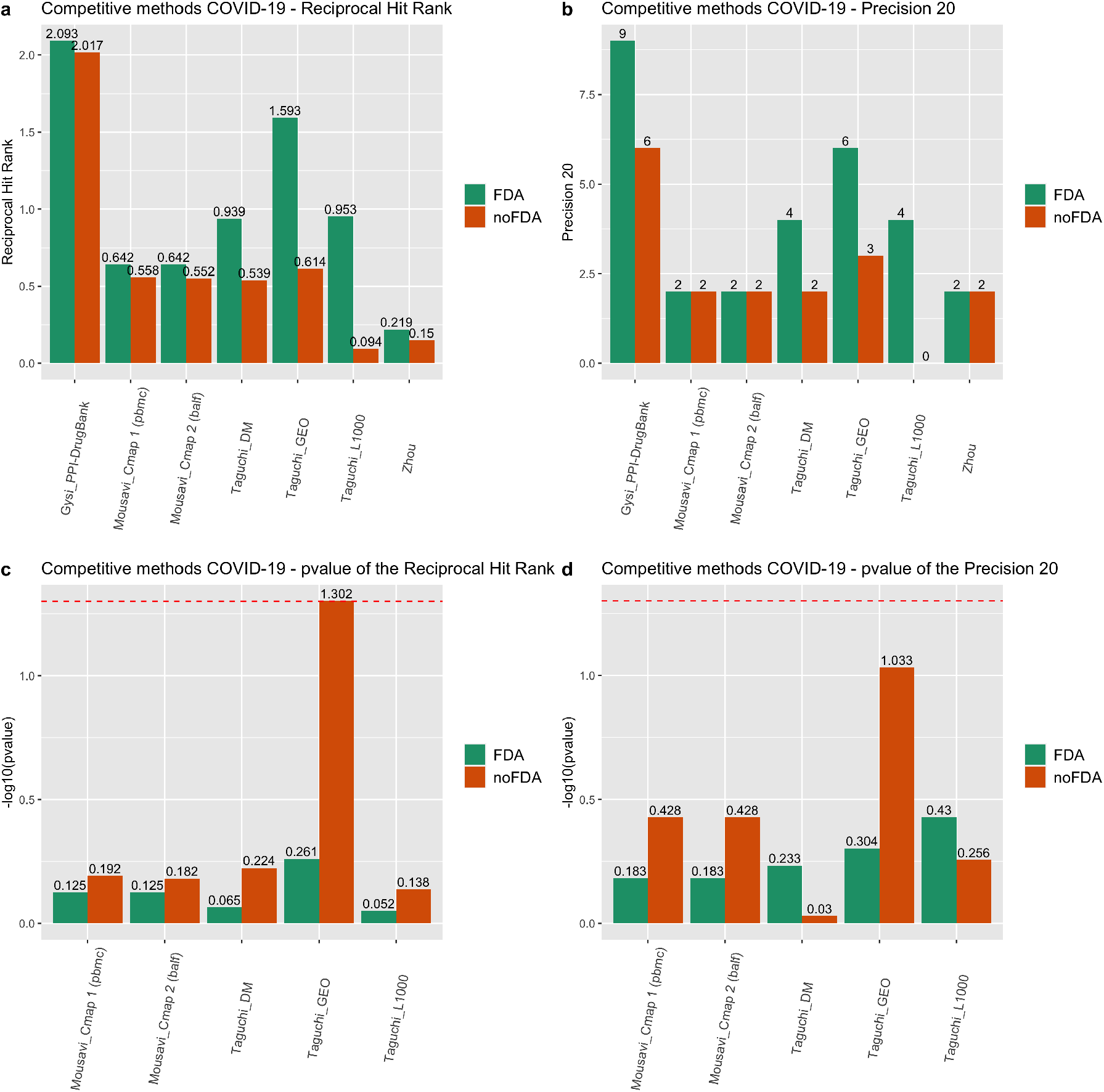
Performance of drug rankings reported in literature for COVID-19 drug repurposing.The dotted red line indicates the 0.05 significance threshold.

Interestingly as shown in Supplementary File S8 no such rankings attain the significance threshold p-value *≤*0.05 in any configuration.

For Gysi et al. 2020 and Zhou et al. 2020, the drug data sets used are different from the ones we use, and the two methods are not based on gene expression data, and it is not possible to determine with confidence the initial pool of drug candidates. For this reason, we do not compute the normalized significance, and we report directly the absolute performance measures. The results of Gysi et al 2020 are in absolute value superiors to those of Zhou et al 2020 and qualitatively comparable to those obtained by DrugMerge on GEO data for precision@20, and on all four Drug datasets for RHR. In particular, Gysi et al 2020 [Gysi et al., 2020] use data on interactions between viral and human proteins in [Gordon et al., 2020] to seed their active subnetwork and finish their listing with the help of expert advice, which we do not use. Gysi et al. 2020 also have a notion of ‘positive’ drug action akin to the one used in our paper.

## 3 Methods

### 3.1 Drug Ranking based on p-value

We use the Core&Peel technique described in [Lucchetta, 2020], [Pellegrini et al., 2016] and 4 other methods (ClustEx [Gu et al., 2010], ModuleDiscoverer [Vlaic et al., 2018], Degas [Ulitsky et al., 2010], and KeyPathwayMiner [Alcaraz et al., 2011]) to produce active subnetworks for a given disease *D*, starting from a list of differentially expressed genes in samples affected by the disease w.r.t. healthy controls *DEG*(*D*) and a gene co-expression network. In [Lucchetta, 2020], we employed the gene co-expression network made available by the DREAM challenge [Choobdar et al., 2019], and DEG for seven different datasets: two inflammatory disease microarray experiments (asthma and rheumatoid arthritis), two cancer RNA-Seq studies (prostate and colorectal cancer), and three COVID-19 datasets called BALF (bronchoalveolar lavage fluid RNA-Seq), PBMC (infected patient peripheral blood mononuclear cells) and COVID19 cells (human adenocarcinomic alveolar basal epithelial A549 cells). In [Lucchetta, 2020], we compared the performance of Core&Peel and the other four algorithms in detecting active subnetworks, resulting in that Core&Peel is a competitive method in that purpose. In this work, we use the five active subnetworks detected by all five methods as inputs of DrugMerge. Let *AN* be any such active network. For a given Drug perturbation datasets (for us: GEO, DrugMatrix, CMAP, and L1000) using the *enrichR* R/Bioconductor package [Kuleshov et al., 2016], we obtain a listing of drugs enriched in *AN* ranked by their adjusted p-value in reverse order (from the smallest to the largest). We cut this list at the adjusted p-value of 0.05, retaining only the significantly enriched drugs.

### 3.2 Drug perturbation databases

To predict the potentially repurposable drugs in each active subnetwork (one for each disease and each method), we used four drug perturbation databases available in Enrichr (https://maayanlab.cloud/Enrichr/#stats). These databases are called ‘Drug_Perturbations_from_GEO_2014’, ‘DrugMatrix’,

‘LINCS_L1000_Chem_Pert_down’, ‘LINCS_L1000_Chem_Pert_up’, ‘Old _CMAP_down’, and ‘Old_CMAP_up’. The GEO- and DrugMatrix-related analysis has been performed in June 2020, that one for L1000 and CMAP has been performed in January 2021. Each database includes several drugs and the corresponding genes perturbed by the drug, distinguishing if these genes are up- or down-regulated. The Drug Perturbations database from GEO was compiled using experiments from GEO [Barrett et al., 2012] where gene expression levels were measured before and after the administration of a drug. This library has drug signatures for several tissues, and species including the *homo sapiens, mus musculus* and *rattus norvegicus*. In total, there are 701 terms and 132 unique drugs (where we do not consider the species, the tissues, and the modularity of the genes). DrugMatrix (DM) [Svoboda et al., 2019], [Ganter et al., 2005] is one of the largest rat toxicogenomics databases and has drug signatures available on liver, heart, kidney, thigh muscle, and primary hepatocytes. In total, it includes 7876 terms and 656 unique drugs. The Connectivity Map (also called CMAP) [Lamb et al., 2006], [Lamb, 2007] is a collection of gene expression data from five different human cells perturbed with many chemicals and genetic reagents. In total, CMAP applied 1309 compounds yielding 6100 profiles. The Connectivity Map project entered a most recent version of CMAP, as part of NIH’s Library of Integrated Network-Based Cellular Signatures (LINCS) program, called LINCS-L1000. It comprises ∼ 5000 genetic perturbations and ∼ 15000 perturbations induced by chemical compounds [Musa et al., 2018] across 98 different cell lines. The two CMAP projects differ also in sequencing platforms. The LINCS-L1000 project replaced the Affymetrix GeneChips used by the original CMAP with Luminex bead arrays [Lim and Pavlidis, 2019], which has been developed to facilitate rapid, flexible, and high-throughput gene expression profiling at lower costs. Using the *enrichR* package, in the L1000 database, we obtained 33132 terms and 4117 compounds.

### 3.3 Filtering of drug ranking

We have noticed that a straightforward application of the p-value based drug ranking includes in top ranking positions some molecules and drugs that are known to be either generally toxic or to induce diseases in animal models. For this reason, we introduce the option of applying two filters to a ranked list of drugs. The first filter is used to retain only drugs that are approved by the FDA (we use a list of drugs downloaded from https://www.accessdata.fda.gov on September 9th 2020, with 6315 entries). For FDA-approved drugs usually adverse effects and toxicity levels are known and probably acceptable in most cases. The second filter retains drugs that by the directionality analysis appear to have a ’positive’ action on the disease, i.e. genes are up/down regulated antagonistic to the disease effect. This list of positive drugs can be obtained with the method described in the next section.

### 3.4 Drug ranking based on drug positivity scores

For this analysis, we exploit the fact that two of the methods we employ to build active subnetworks produce the active sub-network as a union of dense sub-graphs (ego-networks) of a large co-expression network (in detail this co-expression network has been produced for the DREAM challenge on disease module detection [Choobdar et al., 2019]). Since such network is built on the correlation of the expression-vectors over thousands of conditions, a high correlation vector implies that a pair of genes are generally both up or both down regulated at any given time. In a dense ego-network, such tendency is coherently common to all genes in the ego-network. We exploit this phenomenon as follows. For an ego-network that is significantly enriched in genes down-regulated by the disease, we subtract the number of up-regulated genes by the drug from the number of down-regulated genes by the drug. If this number is positive we say that the drug has a *positive* effect of the disease (for this ego-network). We do a symmetric calculation of the case of ego-networks that are significantly enriched in genes up-regulated by the disease. Summing these effects (with their sign) over all the ego-networks comprising the active subnetwork, we can compute the directionality of action of the drug and a magnitude (directionality score). We retain the drugs that have a positive directionality score and we rank them by the magnitude of the score. Note that this scheme relies just on counting the number of up/down regulated genes, not on the magnitude of the expression, as long as it is significant. In a variant of this scheme we can use the size of the ego-networks as a weighting factor. This method is justified by an extension to the ego-networks of the phenomenon Chen et al. [Chen et al., 2017] have noticed that in preclinical models of breast, liver, and colon cancers, reversal of gene expression between drug and disease correlates well with drug efficacy. Furthermore, Chen et al. validated the efficacy of drugs in xenograft models of the diseases.

### 3.5 Merging of Drug lists

Given two or more ranked lists of drugs, we wish to aggregate them into a single list that takes into account the ranking of any drug in each of the lists where it occurs. Following Gysi et al. [Gysi et al., 2020] we use the C-rank routine in [Zitnik et al., 2018b] to perform rank aggregation. C-rank is defined in [Zitnik et al., 2018b] to work with any custom-defined ranking of items^7^. Interestingly this rank aggregation method is oblivious of the origin of the ranking and we can thus easily aggregate lists obtained with different active subnetwork algorithms on different disease-DEG sets. The only caveat is that the lists supplied to c-rank must be of equal length, thus the case of missing drugs must be handled, by fitting the drugs missing from a list at its bottom with a scaled mean score value obtained from the other lists.

### 3.6 Performance evaluation

The performance evaluation aims at assessing how well a predicted ranked list gives priorities in accordance to human expert judgment as codified by disease-treatment pairs extracted from the *Therapeutic Target Database* (TTD) [Chen et al., 2002] for the four benchmark diseases. Since for COVID-19 currently only one drug has gained acceptance as an approved clinical treatment, we resort to the list of drugs for the treatment of COVID-19 undergoing clinical trial as listed in the portal https://www.clinicaltrials.gov/ (at Sept 5th, 2020).

We use two main measures. The first is precision at 20 (precision@20), that is the number of drugs in the first 20 positions of the proposed list of repurposable drugs that are hits in the TTD-derived listing, or, for COVID-19, in the clinicaltrials.gov listing.

The second is the Reciprocal Hit-Rank (RHR) [Deshpande and Karypis, 2004]. For a list *L* of predictions length *N*, let the hits be in positions *p*_1_, *p*_2_, …, *p*_*h*_. We define :

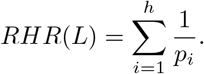

Note that this formula does not imply a fixed length for the lists being compared, and it is fair in comparing both long and short lists. A longer list may have more hits by chance, however it will have positive but diminishing returns for hits in its tail. We prefer not to use ROC-related estimates that may be inaccurate when the hits and miss classes are very unbalanced [Hanczar et al., 2010] [Brown and Patel, 2018].

### 3.7 Statistical significance

Let D be a set of drugs and *H* ⊂ *D* a golden standard for a disease (e.g. the drugs in *D* that are in clinical use for the disease). We generate *n* random permutations uniformly at random *r*_*i*_(*D*), for *i* = 1, .., *n* and we take, for a performance measure *µ*_*H*_ () depending on *H*, the mean *m* and standard deviation *sd* of the sequence *M* = [*µ*_*H*_ (*r*_*i*_(*D*)) for *i* = 1, .., *n*]. For a given drug ranking *R*(*D*), its *zscore* for *µ*_*H*_ () is its deviation from the mean of the above described random distribution normalized by the standard deviation of the random distribution:

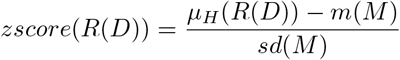

Assuming that the random distribution of the values of *µ*_*H*_ (*r*_*i*_(*D*)) is normal, we obtain the (two sided) p-value by plugging the z-score value into the complementary cumulative distribution function (ccdf) of the normal distribution from the package scipy.stats of SciPy.org.

The number *n* of random permutations is determined as follows. Starting with *n* = 100, 000, we increment *n* in steps of 10, 000, stopping when the value of both *m* and *sd* are stable, with a displacement from the previous iteration less than 0.01.

### 3.8 Drug name and aliases

We handle drug aliases by using the MESH^8^ database. Moreover, we identified commercial drug names with their principal active ingredient. Ambiguous cases were checked by hand against the pharmacological literature and DrugBank records (https://go.drugbank.com/).

### 3.9 CMAP algorithm

To compare the DrugMerge performance in the four benchmark diseases (asthma, rheumatoid arthritis, prostate cancer, and colorectal cancer), we used the *PharmacoGx* [Smirnov et al., 2016] R package, which performs the Connectivity Map (CMAP) analysis [Lamb et al., 2006]. For each disease, we used the differentially expressed genes (DEGs) as gene input to identify drugs with therapeutic potential in each disease. In particular, we used the *downloadPSet* function to download the CMAP database, and the *drugPerturbationSig* function to identify differential gene expressions induced by drug treatment. Finally, the *connectivityScore* function compares drug signatures against disease signatures (DEGs), by assigning connectivity score and p-value for each drug. The connectivity score determines the correlation between the drug and disease signatures and it ranges from *-*1 to 1. Since we are looking for disease treatments or drug repurposing, the CMAP drugs should be anti-correlated with disease signatures. For this reason, we ordered the final CMAP drugs according to the increasing order of the connectivity score (from negative to positive values). We also filtered the CMAP drugs according to p-value < 0.05 and we used both lists with or without p-value filtering. Starting from these lists, we calculated the RHR and precision@20 with z-score and p-values associated. After that, we selected those results with the best values of RHR and compare them with values detected by DrugMerge as reported in Figure 6 and Table S6.

## 4 Discussion

We can identify three main strategies in Drug Repositioning (DR): the first is disease-oriented DR (i.e. find repurposable drugs for a specified disease), the second is drug-oriented DR (finding a disease for a specified drug), the third is generic DR (find any novel drug-disease pair). The approaches may vary significantly across the three types, and also the validation methodology is strongly influenced by the objective of the strategy. In this study we tackle disease-oriented DR, however, we should point out that the base methodology can in principle be adapted easily to the other two strategies, provided sufficient high-quality data is available.

DrugMerge is a proof-of-concept exploring a novel use of disease active subnetworks in the context of drug repositioning aimed at a specific disease. The tests involving four benchmark diseases with drugs in clinical use did confirm the superior performance of the proposed method over the current state of the art (e.g. CMAP) in placing such clinically approved drugs in top ranking positions. In this case, the evaluation criterion is very strict since we were able to guess correctly the efficacy and safety of drugs administered to patients basing our analysis only on transcriptomic data from *in vitro* cell lines and tissues. Interestingly, while efficacy is in principle detectable by examining specific types of cells or tissues primarily affected by a disease, safety is a much broader concept involving all human organs and tissues, thus harder to pinpoint with localized experiments either *in vitro* or *in silico*.

Tests on the data from the COVID-19 disease also show, according to our validation criterion, a performance superior to some of the approaches proposed in the literature, while for some others we notice a broadly equivalent outcome, even when using different approaches and different input data. Results on COVID-19 should be taken with caution, however, as our validation methodology implies that the proposed ranking is able to mimic the best judgment of human experts, which may be based on their prior knowledge and expertise, data available to them, and their assessment of the odds, leading to a shortlisting of such drugs in clinical trials. Thus our validation methodology might indeed be failing us in cases where the disease to be tackled has key features that escape our present general understanding. What can be gained in our exercise is a way to make drug shortlisting for *in vitro* further assessment (and eventual accession to clinical trial) more systematic, while also using relatively cost effective data sources. Besides the global gene co-expression network, which we can assume as background knowledge, our method relies on rather standard and relatively inexpensive RNA-Seq and microarray transcriptomic measurements of differentially expressed genes in infected cells/tissues. Systematization and finding alternatives to pure serendipity (discovery by chance) are key aspects of all *in silico* based drug repositioning efforts.

One bonus of this parsimonious data requirement is that it becomes easier to track the evolution of viral Mechanism of Action and disease etiology as the viral agent mutates, thus in principle the drug ranking, and thus follow up decisions, may change rapidly when transcriptomic data on new viral strains of COVID-19 become available.

Sars-Cov2 is a member of the coronavirus family, responsible also for the Sars-CoV and MERS epidemics. Many studies, in particular at the end of 2019 and beginning of 2020, make up for the paucity of data on Sars-Cov2 by making use of genomic, transcriptomic, proteomic, and pharmacological data relative to Sars-CoV and MERS and other members of the coronavirus family, including those known to be hosted by other animal species.

In our study, we chose not to use data from viruses different from Sars-Cov2 as primary data. This choice comes from two orders of considerations. The first is that data from close relatives of the pathogen may not always be available, and there is thus a general long term advantage attained by methods that rely just on relatively small and focused data. The second issue is that we focus not on the pathogen in itself but on the human transcriptional response to it, which is known to be rather different across Sars-Cov2, Sars-CoV, and MERS epidemics. In this line of reasoning, it makes more sense to highlight what makes human transcriptional response to Sars-Cov2 different from that induced by other coronaviruses, even when the causing pathogens are phylogenetically very close.

We use the list merging procedure to merge lists obtained by several algorithms and several DE gene data sets from different cell lines/tissues but always referred to a single drug data sets, and a single disease. Different drug data sets are often obtained using different initial candidate drugs and use different technologies. Moreover sometimes, due to the different technologies and other experimental settings, even when we restrict the analysis to common drugs, replicability across drug data sets may be low (see e.g. [Lim and Pavlidis, 2019] [Lin et al., 2020] reporting low replicability across CMAP and L1000 data using the CMAP algorithm). Moreover the performance measures we use measure the quality of the whole ranking, and the ranking itself gives a way to compare drugs listed by position. However, based on these measures we refrain from comparing (and thus give a relative order) to two drugs in different rankings obtained with different perturbation drug data sets. Such a global ranking would need further insight and technological bias correction, which we leave for future research.

Drug DE gene data report often measurements for a range of different dosages taken at a range of different time points. For the p-valued ranking, we use the dosage/time point giving the lowest p-value. For rankings based on positivity score we take the average positivity score. These choices are justified for an initial assessment of drug potential to be effective against a disease. Dosage and time points could be taken into account together with available dose-dependent toxicity data in a second level of analysis when additional data is collected in *in vitro* experiments.

For this study, we use all available transcriptomic drug and disease data regardless of the cell lines/tissues/species for which the transcriptomic measurements are taken. This choice increases the robustness of the results in general, however, it might miss some more subtle tissue-specific phenomena [Newman et al., 2020].

On the one hand, it is known that the SARS-Cov2 virus infects several human organs includeing lungs, liver, kidneys, and gut [Mallapaty, 2020], and the cells transcriptional response may in principle be diverse in each tissue. However, Dudley et al.[Dudley et al., 2009] analyzing a large repository of disease-related gene differential expression data conclude that “molecular signature of disease across tissues is overall more prominent than the signature of tissue expression across diseases”. Our analysis of active subnetworks extracted from three COVID-19 data sets (BALF, PBMC, cells) also supports for COVID-19 a general high similarity of gene expression across the sample types. Differences in the extracted active networks are mainly due to the algorithms used.

Also, drugs may act differently in different cell types. For example, Chloroquine has been found to be ineffective in infected human lung cells [Hoffmann et al., 2020], while it was found effective in infected Vero cells (a cell line isolated from kidney epithelial cells extracted from an African green monkey) [Wang et al., 2020a]. Similar tissue-specific differences in drug sensitivity are noticed by Dittmar et al. [Dittmar et al., 2020]. In principle, if a variety of options are available, one may opt for including only the data drawn from biological samples more similar to the actual tissues affected by the disease in patients. In the context of Cancer drug repostioning, a tool like CELLector [Najgebauer et al., 2020] may be beneficial for a systematic cell/tissue pre-selection.

In our study we use a large variety of cellular types for drug perturbation analsis. Drug matrix [Ganter et al., 2005] has drug perturbation signatures available on liver, heart, kidney, thigh muscle and primary hepatocytes (from *rattus norvegicus*). GEO [Barrett et al., 2012] has drug perturbation signatures for several tissues and species including *homo sapiens, mus musculus* and *rattus norvegicus*. CMAP measures drug perturbations in breast cancer epithelial cell line MCF7 (MCF7/ssMCF7), in prostate cancer epithelial cell line PC3, and the nonepithelial lines HL60 (leukemia) and SKMEL5 (melanoma). L1000 collects drug perturbation data from 98 different cellular contexts including human primary cell lines and human cancer cell lines [Lim and Pavlidis, 2019].

For COVID19 we report two drug rankings attaining top performance, one is obtained by merging data from all three tissues used to measure DE genes, therefore it indicates evidence for drugs that may act on a general systemic level across different human organs. This is complemented by the second ranking which is attained specifically for BALF measurements (bronchoalveolar lavage fluid) and thus it may indicate drugs with a specific action in pulmonary tissues.

One advantage of our approach, in contrast with many others, is that we do not have an initial explicit guess for a putative target protein for the drug or a putative target pathway (or mechanism of action). However, once relevant drugs are shortlisted we can analyze their likely mechanism of action, either through the literature or through the use of specialized databases (e.g. DrugPath [Jaundoo and Craddock, 2020]).

In this study, we have used four public Drug related data sets (GEO, DrugMatrix, CMAP, and L1000) representing a sufficient drug basis for our investigation and the validation of the proposed methodology. However, a much larger amount of useful data on drugs is proprietary therefore it is important to devise incentives for sharing these repositories of useful data in a global drug repositioning strategy. As a consequence of the non-complete drug coverage of our data sets we could not assess some drugs whose use in COVID-19 cases has been advocated, such as remdesivir (approved by FDA in October 2020 [Rubin et al., 2020]) or lopinavir (not approved by FDA). Cao et al. [Cao et al., 2020] recently reported the results of a randomized clinical trial involving 199 hospitalized adult patients with severe COVID-19: no benefit was observed with lopinavir–ritonavir treatment over the standard care cohort. Interestingly, we have data from DrugMatrix on ritonavir (not approved by FDA), which fails the positivity score filter, thus our results on ritonavir are consistent with the findings in [Cao et al., 2020].

Comorbidities in patients affected by COVID-19 are important clinical aspects, as it is known that comorbidities are one of the strongest risk factors for such patients, strongly affecting mortality rates [Sanyaolu et al., 2020]. Comorbidities may alter drastically the effect of drugs. The disease gene expression data (for all five diseases) we use in this study do not come from patients with known comorbidities, therefore the results of our study should be interpreted in such a setting. Some authors ([Fiscon et al., 2020]) try to exploit known comorbidities as a positive tool in modeling the relevant features of COVID-19, while other authors, based on protein target analysis, have a more skeptical view ([Gysi et al., 2020]) : “In summary, we find that the SARS-CoV2 targets do not overlap with disease genes associated with any major diseases, indicating that a potential COVID-19 treatment can not be derived from the arsenal of therapies approved for specific diseases.” The effect of concomitant comorbidities on repurposed drugs might be better approached when transcriptomic data from these sub-populations of COVID-19 patients become available.

In this study, we do not treat the issue of combinations of drugs. This is a novel area of research in which network-based approaches have been shown to give novel insight and may help in coping with the combinatorial explosion of drug combinations. However, validated data on drug combinations is limited thus making the task of performance evaluation and method validation more complex w.r.t the analysis for single drugs. We plan to incorporate models of multi-drug/disease interactions in our setting as future research [Cheng et al., 2019].

Drugs can be toxic or cause adverse events. Toxicity can be detected via ad-hoc *in vitro* ed *in vivo* experiments or it can be predicted via *in silico* models [Zhang et al., 2018],[Ganter et al., 2005],[Igarashi et al., 2015],[Alexander-Dann et al., 2018],[Poleksic and Xie, 2019]. Any of these techniques should be applied downstream to the use of DrugMerge for extra safety. DrugMerge uses two forms of filtering to handle adverse effects and toxicity. The first mechanism is the inclusion of drugs having a contrary effect to the disease as measured by the positivity score (which in turn is similar to a mechanism also used by [Gysi et al., 2020]). The second filtering is via the possibility to limit the analysis to FDA-approved drugs, for which toxicity and adverse effects are known and have been considered to be outweighed by the benefits in certain conditions. This mild form of filtering is sufficient to increase the performance in our tests and thus mimic human expert judgment, which includes consideration of toxicity and adverse effects. However, toxicity should be always be reconsidered ex-post to finalize any shortlisting decision.

Since we base our approach on the transcriptomic responses of human cells/tissues to drugs and diseases, the cause of the disease (viral, bacterial, genetic, metabolic, etc..) is not critical for the application of the methodology. Indeed in our study, we applied it to a variety of cases. Asthma is an inflammatory disease of the airways of the lungs having several triggering causes, including exposure to air pollution and allergens. Rheumatoid arthritis (RA) is a long-term autoimmune disorder that primarily affects joints. Cancer is generally associated with alterations of the genetic code in somatic human cells. Finally, COVID-19 is of viral origin. The advantage of generality is balanced by the possibility that a high ranking drug is effective against symptoms or side-effects of the diseases, rather than acting on its main mechanism of action. In one of the drug rankings for COVID-19, we find within the top 20 positions prochlorperazine which has both antiviral activity [Otręba et al., 2020] and is also a broad-spectrum antiemetic contrasting covid19 symptoms like nausea and vomiting [Kow and Hasan, 2020], [Zhang et al., 2020a]. At this level of analysis, DrugMerge does not yet distinguish the different actions a drug might have thus further investigations are needed to untangle complex drug actions.

## 5 Related work

Computational drug repositioning is a burgeoning field of research (see. e.g. [Li et al., 2016], [Levin et al., 2020] [Galindez et al., 2021], [Dotolo et al., 2020], [Pushpakom et al., 2019], [Alaimo and Pulvirenti, 2019], [Lotfi Shahreza et al., 2018], [Xue et al., 2018], [Jarada et al., 2020], [Low et al., 2020]) receiving much attention for its high potential impact on coping with present and future pandemic events by novel pathogens [Zhou et al., 2020c] [Nabirotchkin et al., 2020].

For the four benchmark diseases considered in our study, studies on repurposed drugs for tumors are reported in [Nowak-Sliwinska et al., 2019], [Zhang et al., 2020b], [Turanli et al., 2018], for asthma in [Kruse and Vanijcharoenkarn, 2018], and for rheumatoid arthritis in [Hu et al., 2019], and [Kim et al., 2018].

A survey focusing on repurposing anti-cancer drugs for covid19 is in [Ciliberto et al., 2020]. Computational drug repositioning is gaining much attention during the current covid19 pandemic as reported in [Singh et al., 2020].

Network-based drug repositioning [Barabasi and Oltvai, 2004] is an approach that leverages on building and analyzing various types of biological networks integrating several layers of ’omic’ data [Re and Valentini, 2013]. Also, drug perturbation data bases play a central role in computational drug profiling [Duan et al., 2014], [Lamb et al., 2006], [Subramanian et al., 2017], [Ganter et al., 2005]. Next, we focus on some results in the lines of research that are most relevant for our study.

Taguchi et al. [Taguchi and Turki, 2020] apply an unsupervised method based on tensor decomposition to perform feature extraction of gene expression profiles in multiple lung cancer cell lines infected with severe acute respiratory syndrome coronavirus 2 [Blanco-Melo et al., 2020]. They thus identified, using Enrichr [Kuleshov et al., 2016], drug candidates that significantly altered the expression of the 163 genes selected in the previous phase. Drug perturbation data is collected from GEO, DrugMatrix, L1000, and other repositories.

Ruiz et al. [Ruiz et al., 2020] build a multi-layer disease-gene-drug-pathway network and develop a random-walk-based score that captures indirect effects of drugs on diseases via commonly affected pathways. They validate their approach in a leave-one-out validation on a golden standard of 6000 drug-disease pairs commonly used in clinical practice. The paper, however, does not report on the specific application of this approach to COVID-19.

Fiscon et al. [Fiscon et al., 2020], [Fiscon et al., 2021] use an approach based on computing distances between regions of a large human interactome network affected by a disease (disease targets) and those affected by drugs (drug perturbations). This work relies on disease target similarities between a group of diseases including covid19, SARS-coV, MERS, and others. The final assessment of drug candidates for COVID19 is done with the C-map database.

Gysi et al. [Gysi et al., 2020] define several proximity-based distance functions between covid19 human target proteins (as listed in [Gordon et al., 2020]) and drug protein targets as listed in DrugBank [Wishart et al., 2018], in order to prioritize repurposable drugs. Several refinement taking into account tissue specificity, drug action on gene expression levels (using data from [Blanco-Melo et al., 2020]), comorbidities, and drug toxicity lead to a final list of 81 repurposable drugs for covid19. The main measure of performance chosen is the AUC of the predicted list versus the list of drugs currently employed in clinical trials for the treatment of COVID-19 as listed in https://www.clinicaltrials.gov/.

Our approach to performance evaluation is similar to the one in [Gysi et al., 2020] since we also use drugs in clinical trials as the golden standard for covid19. Thus we are both measuring how well our automated ranking systems come close to the collective wisdom of human experts that shortlisted existing drugs for repurposing on COVID-19 during 2020 based on a large variety of considerations such as available clinical and preclinical data, pharmacological background, guesses mechanism of action. However, we decided to use precision@20 and the Reciprocal Hit Ranking as quantitative measures since they are quite intuitive, more suitable for a ranking problem, and they handle more uniformly lists of candidate drugs that can span from a few dozen drugs to a few thousand.

Zhou et al. [Zhou et al., 2020b] also use proximity-based measures in biological networks to find a list of 16 repurposable drugs. It should be noticed that Zhou et al. use data collected from the family of human coronaviruses (HCoVs) to build the network, thus relying heavily on evolutionary conservation of relevant coding parts of the viral genome across the species in this family and on phylogenetic considerations.

Sadegh et al. [Sadegh et al., 2020] use the algorithm KeyPathwayMiner [Alcaraz et al., 2011] to define the active subnetwork in an integrated COVID-19 resource network from CoronaVirus Explorer CoVex resource (https://exbio.wzw.tum.de/covex/). The main contribution of [Alcaraz et al., 2011] is a resource (CoVex) that can be used interactively in many different scenarios and modalities to explore disease-gene-drug relationships for COVID-19.

Mall et al. [Mall et al., 2020] use deep learning induced vector embedding of drugs and viral proteins to predict drug-viral protein activity and propose a short list of 15 drugs potentially useful against COVID-19.

A second very popular approach uses knowledge of the 3-dimensional molecular configuration of proteins and drugs to simulate, via docking or molecular dynamics, the most promising drug-target bindings. Most research in docking-based drug repositioning starts with an assessment of one (or a few) protein acting as potential drug targets [Chellapandi and Saranya, 2020].

Seo et al. [Seo et al., 2020] take three dimensional structure of the main protease (Mpro) of SARS-CoV-2 as a target, and use docking simulations on a supercomputer to evaluate binding affinity between Mpro and drug candidates listed in the SWEETLEAD library and the ChEMBL database (19,168 molecules), thus shortlisting 43 drugs, which, after molecular dynamics simulations, are then reduced to 8. A similar docking-based screening is also used in [Gimeno et al., 2020], [Shah et al., 2020], [Elmezayen et al., 2020], [Wang, 2020], [Trezza et al., 2020], and [Mahapatra et al., 2020].

Besides network-based and docking-based approaches there is a variety of other principles and approaches that have been used both *in silico* and *in vitro*: Luminescent cell viability assay and cell imaging [Riva et al., 2020a] [Riva et al., 2020b], morphological cell profiling [Mirabelli et al., 2020], co-morbidity [Luna et al., 2020], deep learning [Zeng et al., 2020], network proximity based [Stolfi et al., 2020], quantum mechanical scoring [Cavasotto and Filippo, 2020], multi-objective optimization [Chen et al., 2020], bipartite network projections [Re and Valentini, 2013], matrix factorization [Huang et al., 2020], machine learning [Loucera et al., 2020], network node scoring [Nam et al., 2020] [Ge et al., 2020], semi-supervised learning [Iorio et al., 2015], variants of gene set enrichment analysis (GSEA) [Fang et al., 2021], multi-omic integration [Tomazou et al., 2021].

A more direct experimental approach has been described in [Gordon et al., 2020] and [Tutuncuoglu et al., 2020]. Gordon et al. [Gordon et al., 2020] performed a direct proteomic assay to uncover 332 human proteins potentially interacting with 26 SARS-Cov2 proteins. Sixty-nine drugs targeting these proteins are then shortlisted as repurposable drugs for covid19, using a mix of chemoinformatic searches and expert advice. Note that in this setting a priority ranking is not provided. A main theme along with drug ranking is that of drug combinations [Muratov and Zakharov, 2020]. Often drug combinations offer lower toxicity and more effective disease treatment. Most drug combinations currently under consideration result from ad-hoc considerations, as screening drug combinations automatically incurs easily in a combinatorial explosion of cases to be considered. A few network-based principled approaches have been proposed and may be integrated into DrugMerge [Artigas et al., 2020], [Zhou et al., 2020b], [Bobrowski et al., 2020], [Bobrowski et al., 2021], [Jin et al., 2020], [Cheng et al., 2019], and [Zitnik et al., 2018a].

While the final hallmark of success is the identification of a repurposed drug that can pass successfully all stages of drug approval for clinical use (see e.g. [Edwards, 2020]), it is important to be able to perform *in silico* validation on intermediate results. A recent study by Brown and Patel [Brown and Patel, 2018] gives a critical assessment of the options for *in silico* drug validations, assessing the weak and strong points of each strategy.

## 6 Conclusions

In this study, we describe DrugMerge, a methodology for ranking repurposable drugs where we rank drugs by their ability to affect collections of disease active subnetworks (computed by an ensemble of stat-of-the-art algorithms) and contrast the perturbation induced by the disease in such sub-networks. The rationale behind our approach is a model in which drugs and diseases interact indirectly by their mutual action on the sub-networks rather than directly by affecting the same target proteins. Moreover, we are able to leverage on an ensemble of different algorithms for computing active subnetworks, and then merge the resulting rankings, thus increasing the robustness of the methodology.

Results of four benchmark diseases are encouraging since we could find in all four cases drugs in clinical use (thus both safe and effective) in first ranking positions, and several other drugs in clinical use in high ranking positions. These results imply that DrugMerge when provided with input data from transcriptional assays, has the potential for coming close to expert human judgment, and guess drugs in actual clinical usage. For COVID-19 we measure how close we come to guessing drugs that have been shortlisted for clinical trials according to the FDA records. For most drugs in high ranking positions but not on clinical trials we have found several supporting evidence in the literature or by direct assessment of their likely Mechanism of Action, that makes their indication by DrugMerge biologically plausible.

DrugMerge should be considered as a proof-of-concept and its predictions should be considered as indications for shortlisting drugs for further *in vitro* analysis rather than absolute predictions. In particular, aspects like dosage, tissue specificity, and a more refined analysis of possible adverse side effects will need to be tackled further in developing DrugMerge towards a full-fledged product.

Prioritizing drugs for *in vitro* testing and eventual accession to clinical trials is valuable for decision makers in a pandemic situation. Lack of prioritization in drug repurposing efforts has led to some drugs being over-researched, early in the pandemic, even based on dubious evidence (e.g. hydroxycholoroquine in COVID-19). Moreover having many small clinical trials has the effect of reducing the pool of eligible patients available for larger and more promising trials^9^.

A tool such as DrugMerge aiming at drug ranking might be used in conjunction with an activity of data collection that targets specific sub-populations of patients. For example, most trials to date for repurposed drugs in COVID-19 have focused on hospitalized patients, even though prevention might be more valuable and effective if initiated earlier in disease progression. Also transcriptomic data from patients affected by frequent comorbidities might help towards the study of drugs specifically selected for sub-populations at higher risk of mortality [Stephenson et al., 2021].

This paper focuses on the preclinical considerations that may indicate a drug to be shortlisted for further preclinical *in vitro* and *in vivo* testing, leading to eventual clinical trial. Naturally, all evidences, which may conceivably be noisy and often contradictory, should be gathered and considered before pushing a drug further down the pipeline.

## Data Availability

Primary data used in this study is available in public repositories. Intermediate results processing
software and scripts will be made available on GitHub.

## 8 Data Availability

Primary data used in this study is available in public repositories. Intermediate results, processing software and scripts will be made available on GitHub.

## 10 Author’s contributions

M.P. designed the study. M.L. performed the analyses. M.P. and M.L. interpreted the results and wrote the manuscript. All authors reviewed the manuscript.

## A Supplementary files

S1 **DrugMerge-asthma-table1.csv**. The table reports the ranking function, the filter, drug perturbation data set, algorithmic configuration attaining best performance on the asthma DEG dataset. It is reported the reverse hits ranking and precision at 20, with corresponding z-scores and p-values. The csv file uses ‘;’ as column separator.

S2 **DrugMerge-arthritis-table1.csv** Table similar to S1 for Rheumatoid Arthritis DEG dataset.

S3 **DrugMerge-colorectal-cancer-table1.csv** Table similar to S1 for Colorectal Cancer DEG dataset.

S4 **DrugMerge-prostate-cancer-table1.csv** Table similar to S1 for Prostatate Cancer DEG dataset.

S5 **DrugMerge-covid19-table4.csv** Table similar to S1 for Covid19 DEG datasets.

S5.1 **Table-algorithm-alias.cvs** Table mapping synthetic algorithms identifiers in Figures 7 and 8 with algorithmic configurations in Table S5.

S6 **Summary-four-diseases-CMAPalg-vs-DrugMerge.xlsx** Table reporting comparative performance of CMAP and DrugMerge on four benchmark data sets (Asthma, Rheumatoid Arthritis, Prostate cancer, and Colorectal cancer). It is reported the reverse hits ranking and precision at 20, with corresponding z-scores and p-values.

S7 **Drug-lists-final.xlsx** Drug rankings with best rhr pvalue performance for DrugMerge on four benchmark data sets (Asthma, Rheumatoid Arthritis, Prostate cancer, and Colorectal cancer), and covid19 (both with FDA approved only drugs, or all drugs).

S8 **Summary-table-comparative-results.xlsx** Table reporting comparative performance of several published drug rankings on covid19. It is reported the reverse hits ranking and precision at 20, with corresponding z-scores and p-values.

## Footnotes

1 https://www.mayoclinic.org/diseases-conditions/asthma/in-depth/asthma-medications/art-20045557

2 In clinical trial https://clinicaltrials.gov/ct2/show/NCT00153270

3 In clinical trial https://clinicaltrials.gov/ct2/show/NCT00379600

4 https://www.drugs.com/monograph/hydrocortisone-systemic.html#ra

5 In clinical trial https://clinicaltrials.gov/ct2/show/NCT00961571

6 In clinical trial https://clinicaltrials.gov/ct2/show/NCT00276549

7 Thus we do not make use of the graph-based community ranking scores for which c-rank was originally designed, instead we use it here as a generic rank aggregation method

8 https://www.nlm.nih.gov/mesh/

9 Generic Drug Repurposing for COVID-19 and Beyond. Susan Athey, Rena Conti, Richard Frank, and Jonathan Gruber Institute for Health System Innovation & Policy. July2020. https://www.gsb.stanford.edu/faculty-research/publications/generic-drug-repurposing-covid-19-beyond

